# Drug response profiling guides precision therapy in relapsed and refractory childhood acute lymphoblastic leukemia

**DOI:** 10.64898/2026.04.08.26350164

**Authors:** Fabio D. Steffen, Andrej Lissat, Julia Alten, Andras Kriston, Nastassja Scheidegger, Cornelia Eckert, Nicole Bodmer, Larissa Schori, Selina Schühle, Arianna Arpagaus, Silvia Gutnik, Despoina Manioti, Noemia Bruderer, Aida Zeckanovic, Imre Västrik, Gergely Nyiri, Ferenc Kovacs, Bodil E. Thorhauge Als-Nielsen, Andishe Attarbaschi, Annika Rademacher, Sarah Elitzur, Elad Jacoby, Barbara De Moerloose, Petter Svenberg, Philip Ancliff, Lucie Sramkova, Barbara Buldini, Adriana Balduzzi, Judith M. Boer, Monika Mielcarek, Francesco Ceppi, Marc Ansari, Jörg Halter, Kjeld Schmiegelow, Franco Locatelli, Francesca DelBufalo, Martin Stanulla, Andreas E. Kulozik, Martin Schrappe, Pierre Rohrlich, Hélène Cavé, André Baruchel, Arend von Stackelberg, Gunnar Cario, Peter Horvath, Beat C. Bornhauser, Jean-Pierre Bourquin

## Abstract

Children with relapsed or refractory acute lymphoblastic leukemia (ALL) require more effective and less toxic therapies. We established a prospective, multicenter Drug Response Profiling (DRP) registry (*NCT06550102*) integrating functional testing into precision-guided treatment. DRP was performed for 340 patients from 17 European countries with a turn-around time of two-weeks. Image-based drug screening with over 135’000 unique perturbations revealed a heterogeneous landscape of *ex vivo* responses to 88 drugs on average. Ranking drug responses across the patient cohort defined individual drug fingerprints, identifying “DRP twins” by similarity in sensitivity and resistance independent of genetic ALL subtypes. Of 239 high-risk patients with follow-up, DRP-informed interventions were reported for 63 patients (26%). Patients received combination therapies based on venetoclax, tyrosine kinase inhibitors, trametinib, bortezomib or selinexor, resulting in objective clinical responses in 43 cases (68%). Precision-guided treatments allowed bridging to cellular therapies in 42 patients among whom 28 (67%) were still alive with a median follow-up of 21 months after DRP (IQR: 14.7-26.6 months). Top responders to venetoclax, ranked within the first tertile of the cohort, had superior 1-year event-survival compared to venetoclax non-responders (0.57 [95% CI, 0.39-0.85] vs. 0.25 [95% CI, 0.11-0.58]). Collectively, these findings demonstrate the feasibility and clinical relevance of functional profiling within an international network. This scalable framework enables individualized therapy selection for enrolment in adaptive precision trials for high-risk pediatric ALL.

## Main

Despite advances in immunotherapy, treatment of children and adolescents with high-risk, relapsed or refractory acute lymphoblastic leukemia (r/r ALL) remains challenging^1–3^. The biological heterogeneity of ALL^4,5^ complicates a “one-size-fits-all” approach based on empirically derived chemo- and immunotherapy protocols, underscoring the need for individualized treatment strategies. This is particularly evident in relapsed T-ALL, where prognosis remains poor^6^.

Genomic profiling enables molecular classification for prognostic and therapeutic stratification. However, robust molecular biomarkers to predict patient responses are rare. Consequently, complementary functional assays are required to assess drug response *ex vivo* and reveal targetable vulnerabilities of the cancer cells^7^. The therapeutic promise of precision medicine approaches is illustrated by the successful integration of tyrosine kinase inhibitors with blinatumomab in ABL-class ALL^8^, BCL2 inhibitors with chemotherapy in r/r ALL and AML^9^ and emerging menin inhibitors in HOXA-activated subtypes^10^. Especially as a bridging regimen, precision-guided treatment (PGT) offers the chance for deeper remission rates, lower toxicity and improved outcomes upon consolidation with CD19- and CD7-directed immunotherapy or hematopoietic stem-cell transplantation (HSCT)^11–13^.

Pediatric genomic profiling programs have primarily focused on solid tumors, demonstrating PGT decisions in 15-30% of patients^14–18^. In the Australian ZERO Childhood Cancer program, 29% of patients receiving PGT experienced clinical responses, with superior 2-year progression-free survival (PFS) compared patients without matched therapy. Early integration of PGT into treatment improved 2-year PFS to 42% versus 12% for later interventions^17^, providing evidence that timely, biologically informed treatment adaptation can translate into clinical benefit.

Functional precision medicine (FPM) and in particular high-throughput drug response profiling (DRP) complements genomics by rapidly capturing integrated, phenotypic dependencies of the cancer cells^7,19–24^. In adult acute myeloid leukemia^25–28^, multiple myeloma^29^, and other aggressive blood cancers^30,31^, DRP has yielded recommendations for targeted treatment in the majority of patients. However, only about a third ultimately received PGT due to early clinical deterioration, limited access to drugs, or regulatory barriers^25,26,30,31^.

In treated subgroups, matched therapies induced measurable responses and improved survival, providing a rational for the next generation of precision trials^32^. In pediatric AML and ALL, recent functional studies have outlined the diversity in drug response across leukemia subtypes^19,23,33–35^. To effectively translate DRP into clinical decision making, rapid turn-around and robust drug scoring are critical. To date, the experience of DRP in a real-time, co-clinical setting, has been limited.

Here, we report the results of the first prospective, international, multicenter implementation of DRP for childhood r/r ALL. We profiled 340 patients from 17 European countries within a federated academic network to demonstrate feasibility and clinical utility of FPM for pediatric ALL. Using automated microscopy, we measured single-cell responses to a panel of 88 drugs on average, generating patient-specific drug sensitivity fingerprints. Actionable results were returned to the physicians within two weeks, informing personalized treatment decisions. DRP confirmed reciprocal dependencies between BCL2- and kinase-driven signaling in T-ALL and revealed subtype-independent drug sensitivities. Importantly, DRP guided the individualized selection of bridging elements for further consolidation with HSCT and CAR-T, improving overall and event-free survival. Together, these findings establish DRP as a scalable functional precision medicine platform for pediatric ALL, bridging the turnaround and implementation gaps of genomic-only approaches and complementing static molecular analyses within international tumor boards.

## Results

### Real-time functional profiling is feasible across federated European centers

We established a real-time, multicenter Drug Response Profiling (DRP) registry (*NCT06550102*) to enable centralized *ex vivo* functional testing as a complementary diagnostic for high-risk ALL. Between January 2022 and December 2023, 156 patients with primary resistant, relapsed, or refractory disease were enrolled (**Table 1**). Bone marrow (BM) aspirates or peripheral blood (PB) samples were tested in mesenchymal stromal cell co-cultures using a high-throughput, multi-well drug screening platform (**Fig. 1a**)^19^. After 72 hours of drug exposure, viable cells were identified by automated fluorescence microscopy and classified by lineage with a supervised machine learning model (**Fig. 1b**). Dose responses were fitted to a log-logistic model and integrated across all samples to generate drug sensitivity scores (**Fig. 1c/d**). Results were communicated to treating physicians via national and international leukemia boards, and clinical follow-up was recorded in the registry (**Fig. 1e**).

**Table 1.** DRP registry cohort description (*n* = 156 patients). The full cohort (*n* = 340 patients) is described in Extended Data Table 1.

| Patient characteristics | N = 156 |
| --- | --- |
| **Sex** |  |
| Male | 96 (62%) |
| Female | 60 (38%) |
| **Age group** |  |
| Infants (<1 year) | 7 (4%) |
| Children (1-12 years) | 87 (56%) |
| Adolescents (13-17 years) | 45 (29%) |
| Young adults (18-24 years) | 5 (3%) |
| Adults (>25 years) | 12 (8%) |
| **Diagnosis** |  |
| BCP-ALL | 88 (56%) |
| T-ALL | 57 (37%) |
| T-LBL | 3 (2%) |
| MPAL | 5 (3%) |
| AUL | 3 (2%) |
| **Disease stage** |  |
| Initial diagnosis | 59 (38%) |
| 1st relapse | 73 (47%) |
| Higher relapse | 24 (15%) |
| **Risk** |  |
| HR (Initial diagnosis) | 59 (38%) |
| HR (1st/higher relapse) | 83 (53%) |
| SR (1st relapse) | 14 (9%) |
| **Subtype<sup>1</sup>** |  |
| TCF3::HLF | 4 (3%) |
| TCF3::PBX1 | 4 (3%) |
| KMT2A | 15 (10%) |
| ETV6::RUNX1 | 7 (4%) |
| BCR::ABL1 | 6 (4%) |
| Ph-like | 5 (3%) |
| PICALM::MLLT10 | 4 (3%) |
| NUP214::ABL1 | 3 (2%) |
| ZNF384 | 4 (3%) |
| Hyperdiploidy | 8 (5%) |
| Hypodiploidy | 2 (1%) |
| iAMP21 | 4 (3%) |
| MEF2D | 2 (1%) |
| PAX5alt | 7 (4%) |

| **Patient characteristics** | **N = 156** |
| --- | --- |
| B-Other | 20 (13%) |
| Unspecified BCP-ALL | 7 (4%) |
| Unspecified T-ALL | 46 (29%) |
| Unspecified MPAL/AUL/T-LBL | 8 (5%) |
| **ETP status** |  |
| non-ETP | 49 |
| ETP | 8 |

<sup>†</sup> As reported by the treating physician.

**Fig 1.**
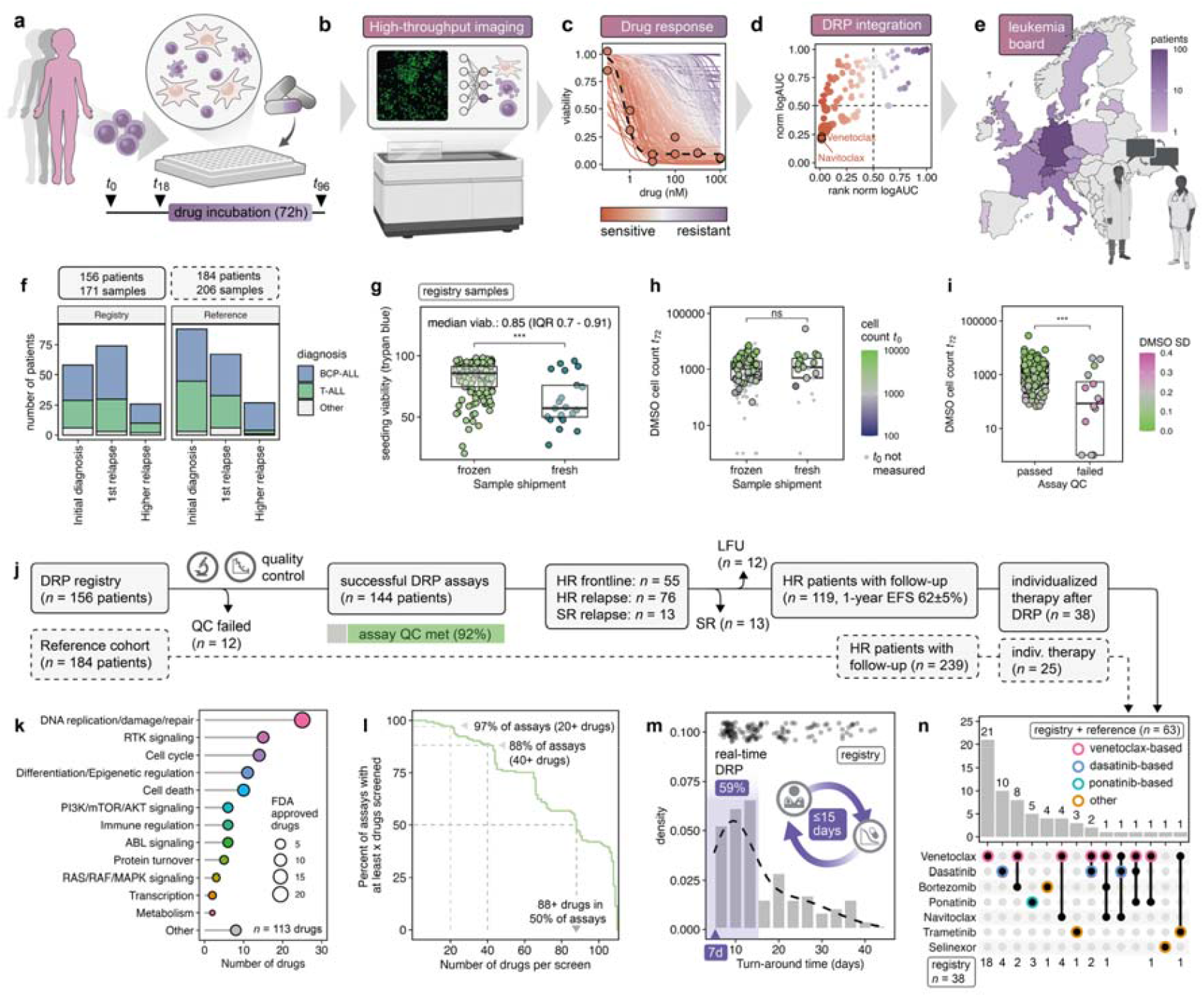
Feasibility of real-time drug response profiling for relapsed/refractory pediatric acute leukemia in a multicenter international setting. **a** Sample collection and drug screening workflow. **b** Automated microscopy and cell classification. **c** Dose response curve fitting. **d** Ranking of drug responses in the patient cohort. **e** Report of DRP results to the treating physician team in an European consortium. **f** Proportions of patients stratified by diagnosis and disease stage for the registry and reference cohort. **g** Viability of cryopreserved and fresh samples before plating the cells, measured by trypan blue. **h** Viable cell counts in vehicle controls determined by CyQuant prior to drug exposure (*t*_0_) and at the endpoint (*t*_72_). **i** DMSO cell count and variation determines assay quality. **j** Consort diagram of patient cohorts including assay success rates, risk stratification, follow-up and individualized treatments after DRP. **k** Drug library categorized by mechanism of action. **l** Proportion of assays for which any given number of drugs was screened. **m** Distribution of turn-around times in the registry cohort (days from sample receipt to report). **n** Numbers of patients treated with one or multiple targeted agents for which DRP information was provided.

The registry cohort comprised pediatric (*n* = 94), adolescent and young adult (AYA, *n* = 50), and adult (*n* = 12) patients (median age = 10 years, IQR 6–15; **Table 1**). Diagnoses included BCP-ALL (*n* = 88, 56%), T-ALL/T-LBL (*n* = 60, 39%), and mixed-phenotype or acute undifferentiated leukemia (MPAL/AUL, *n* = 8, 5%), enrolled at initial diagnosis (*n* = 59, 38%), first relapse (*n* = 73, 47%), or later relapse (*n* = 24, 15%; **Fig. 1f**). As an independent reference cohort, drug response profiles from 184 additional patients tested between 2016 and 2021 in add-on studies to phase II/III trials and compassionate-use programs were included (**Extended Data Table 1**).

To standardize logistics, DRP was performed predominately on viably frozen samples minimizing transit-related variability and increasing assay robustness. In the registry cohort, median viability at seeding was 0.85 (IQR 0.7-0.91). Frozen samples (*n*=149) retained higher viability post-thaw than fresh samples (*n*=22) shipped at ambient temperature **(Fig. 1g)**. However, absolute viable cell counts measured at the endpoint of the assay (*t*_72_) did not differ. (**Fig. 1h**) Baseline survival in vehicle controls (dimethyl sulfoxide, DMSO) was the principal determinant of assay success (**Fig. i**). Quality control (QC) metrics included variation in cell numbers of vehicle conditions (DMSO σ<0.25) and a detection threshold of 50 viable cells in DMSO wells. Furthermore, each plate contained idarubicin as a positive control, producing a median Z′-factor of 0.60 (IQR 0.32–0.74) and confirming reliable effect sizes (**Extended Data Fig. 1**). Overall, QC requirements were met in 143 of 157 patients (91%, **Fig. 1j**).

The number of tested drugs from the 113-compound library (**Fig. 1k**) depended on cell yield. About 18 million viable shipped cells were needed for a full panel screen in duplicate six-point dilutions. At least 40 drugs could be profiled in 88% of cases (**Fig. 1l**), prioritizing subtype-relevant compounds when material was limited. For patients with urgent clinical needs, reports were returned to treating physicians within 7–15 days (**Fig. 1m**), allowing inclusion in precision tumor board discussions alongside molecular profiling^14^.

Of 118 followed-up high-risk (HR) patients in the registry, 38 received a targeted treatment informed by DRP results (32%, **Fig. 1n**). Across both registry and reference cohorts, 239 HR patients had documented follow-up, of which 63 patients received pathway-directed inhibitors after DRP. Regimens were based on venetoclax (*n*=38, including combinations with bortezomib and navitoclax), dasatinib (*n*=11), ponatinib (*n*=5), trametinib (*n*=4) and selinexor (*n*=1). These data demonstrate the feasibility and clinical applicability of centralized DRP within an international network, supporting its use to guide individualized therapy selection.

### Single-cell DRP defines coherent pharmacological activities across functional classes

To enable clinical implementation, we standardized DRP metrics to objectively quantify drug responses across patients. The image-based DRP assay measured leukemia cell viability from unsorted BM samples at single-cell resolution (**Fig. 2a**). Cell identity and viability were predicted using a supervised classifier trained on image intensity and segmentation features. Normalization to vehicle controls produced log-logistic dose-response curves and patient-specific activity metrics, including logarithmic half-maximal effective concentration (logEC_50_) and integrated area under the curve (logAUC, **Fig. 2b**). Collectively, we measured more than 135’000 unique drug perturbations over multi-log concentration gradients, capturing inter-patient variability more reliably than with fixed-dose exposures.

**Fig 2.**
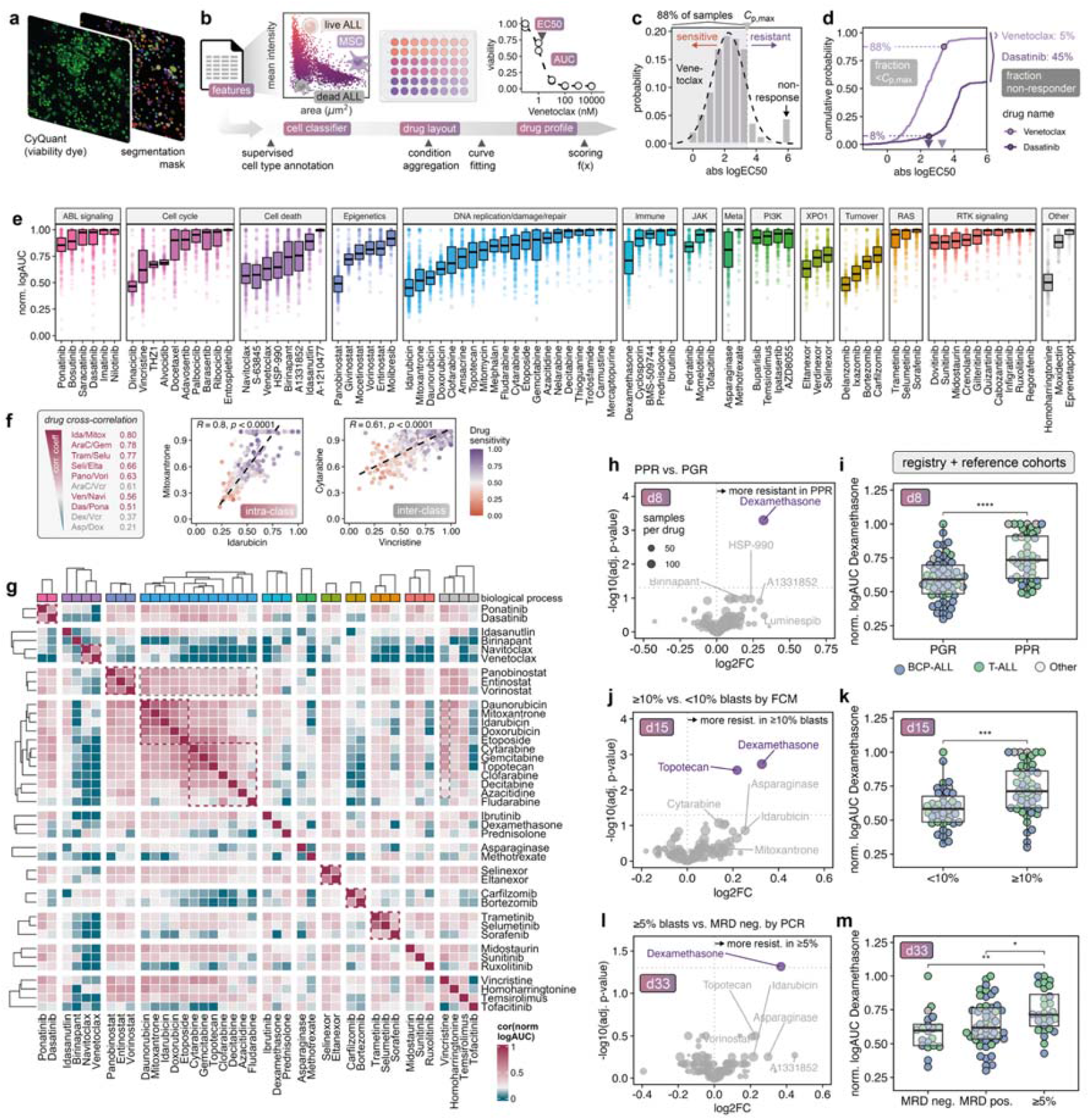
Quantification of recurrent drug activities across pharmacological classes by image-based DRP. **a** Imaging of viable leukemia cells using automated fluorescence microscopy. **b** Cell segmentation and classification to measure drug responses. **c** Normal distribution of absolute logEC50 for venetoclax with 88% of samples displaying an absolute logEC50 below *C*_p_,_max_. **d** Bin-free cumulative distributions of absolute logEC_50_ for venetoclax and dasatinib. Relative proportions of patients below the maximal plasma concentration and fraction of non-responders are indicated. **e** Drug activities represented as normalized, log-transformed AUCs for drugs that have been tested in at least 50% of all assays (88 drugs). **f** Intra-class (red) and inter-class (gray) cross-correlations of drugs with common or distinct molecular mechanisms. **g** Spearman pairwise correlation map of the 40 most frequent drugs across registry and reference cohort shows clusters of drugs with similar sensitivity profiles. **h-m** Volcano plots comparing poor and good responders to the cytoreductive prednisone prephase in firstline treatment (PPR vs. PGR), FCM-MRD response at day 15 (≥10% vs. <10% blasts in BM) and PCR-MRD response at day 33 (≥5% blasts in BM vs. MRD negativity). Corresponding boxplots showing *ex vivo* dexamethasone sensitivity as a function of the clinical response to the steroid prephase (d8), the first week of induction (d15) and end-of-induction (d33). Group comparisons with Student’s t-test.

Distinct activity ranges characterized specific compounds classes (**Fig. 2c-e**). Anthracyclines, proteasome inhibitors, and selected CDK or HDAC inhibitors were highly potent in a narrow nanomolar range, whereas asparaginase, cytarabine, dexamethasone, and BCL2 or XPO1 inhibitors displayed broader activity. Drug efficacy was quantified relative to the sensitivity distribution of all patients. Curated pediatric plasma levels (*C*_p,max_)^36^ contextualized *ex vivo* concentrations with achievable doses in patients. For several drugs, including glucocorticoids and asparaginase, subpopulations of resistant leukemic cells persisted beyond physiological exposure ranges, pointing at possible resistance (**Extended Data Fig. 2**).

Targeted inhibitors often exhibited bimodal response distributions with varying proportions of patients responding to the drugs (**Extended Data Fig. 2** and **Extended Data Fig. 3**), depending on target availability and engagement. Compounds inhibiting tyrosine kinases, PI3K/AKT/mTOR and RAS/MAPK pathways were active in the upper nanomolar range. Certain drugs rely on metabolic conversion before reaching the active form. Accordingly, mercaptopurine which requires additional enzymatic activation steps compared to thioguanine, was less cytotoxic *ex vivo*, consistent with its pharmacokinetics.

Pairwise correlation of drug sensitivities confirmed the biological coherence of DRP signals (**Fig. 2f/g** and **Extended Data Fig. 4**). Strong intra-class concordance was observed among topoisomerase inhibitors (idarubicin–mitoxantrone, Spearman’s coefficient *R*=0.80), nucleoside analogs (gemcitabine–cytarabine, *R*=0.78), MEK inhibitors (trametinib– selumetinib, *R*=0.77), XPO1 inhibitors (selinexor–eltanexor, *R*=0.66), and HDAC inhibitors (panobinostat–vorinostat, *R*=0.63). BCL2-family inhibitors (venetoclax–navitoclax, *R*=0.56) and ABL1 inhibitors (dasatinib–ponatinib, *R*=0.51) displayed moderate correlation, reflecting target selectivity differences.

Across drug classes, correlations between standard induction agents varied from low (asparaginase–doxorubicin, *R*=0.21) to intermediate (dexamethasone–vincristine, *R*=0.37). Conversely, drugs used in sequential phases of the protocol showed higher concordance (cytarabine–vincristine, *R*=0.61). These findings suggest that the efficacy of multidrug regimens in ALL arises primarily from additive and complementary effects. According to this model^37,38^, drugs acting independently with low or intermediate correlations increase the likelihood of response in heterogeneous cohorts, conferring clinical benefit for a higher fraction of patients.

### *Ex vivo* DRP reflects heterogeneity in clinical responses to frontline chemotherapy

We next compared *ex vivo* drug phenotypes to clinical response during frontline ALL therapy in the European AIEOP-BFM 2017 study. Poor prednisone pre-phase responders (PPR, ≥1000 blast/μL in peripheral blood) were more resistant to dexamethasone, topotecan, birinapant, and HSP-990 inhibition than prednisone good responders (PGR), indicating shared defects in apoptotic and stress-response pathways (**Fig. 2h/i**). Minimal residual disease (MRD by flow-cytometry) levels ≥10% at day 15 of induction therapy correlated with reduced *ex vivo* sensitivity to dexamethasone, anthracyclines, asparaginase, and pyrimidine analogs (<10%, **Fig. 2j/k**). At end-of-induction, patients with persistent disease (≥5% blasts by morphology or MRD) retained more resistant DRP signatures to glucocorticoids and anthracyclines compared to MRD-negative cases (**Fig. 2l/m**). Dexamethasone resistance was particularly enriched in T-ALL, consistent with seminal observations made decades ago^39^. More recently, *ex vivo* resistance to cytarabine, glucocorticoids, and doxorubicin was reported to be independently associated with outcome^40^. These results indicate that DRP recapitulates clinically relevant patterns of treatment resistance and aligns with independent MRD-based measures of therapeutic response.

### Cohort-ranked sensitivities define individual drug fingerprints

To compare patient samples, we transformed normalized AUC values into rank-ordered fingerprints (**Fig. 3a**/**b**). The drug fingerprint is composed of individual drug ranks, each weighted by the variance of the drug in the cohort, thereby allocating more weight to compounds with larger effect sizes between patients (**Fig. 3c-e**). Amongst the most distinguishing drugs were vincristine, asparaginase, gemcitabine, dexamethasone and BCL2 family inhibitors. Unsupervised clustering of these fingerprints identified patient groups with distinctive clusters of resistance or sensitivity (**Fig. 3f**).

**Fig 3.**
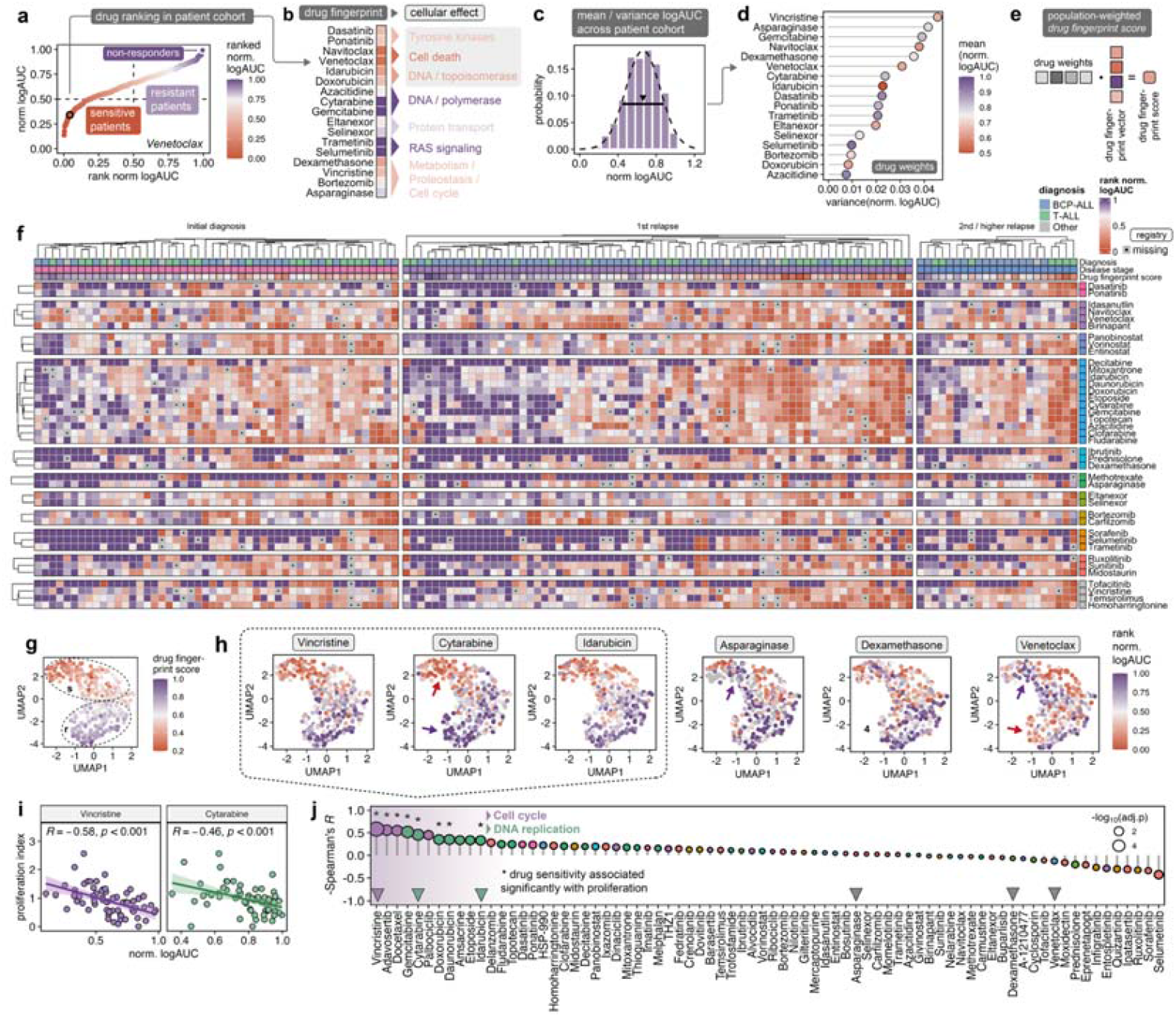
Landscape of drug sensitivities in pediatric ALL across disease stages. **a** Ranking of logAUC values over the combined registry and reference cohort, exemplified for venetoclax. **b** the concatenated drug ranks define a patient’s drug fingerprint represented as a column vector. **c** Distribution of normalized logAUC values. **d** The variance of the distribution assigns a weight to each drug. **e** Calculation of the drug fingerprint score as a weighted mean of drug ranks. **f** Collection of drug fingerprints in the registry cohort clustered separately by disease stage. **g/h** UMAP projection of patients from the registry and reference cohort, color-coded by the drug fingerprint score or individual drug ranks, highlighting differential drug sensitivities in patient subgroups (red and violet arrows). **i** Correlation of drug sensitivity and sample proliferation for two cell cycle dependent compounds. **j** Waterfall plot of Spearman coefficients between the normalized logAUC and the proliferation index color-coded by functional class.

Upon combining BCP- and T-ALL and reducing drug dimensions by uniform manifold approximation and projection (UMAP), we observed a polarization into *ex vivo* good and poor responders (**Fig. 3g**). The embedding highlights inter-patient differences among standard induction agents (**Fig. 3h**). The pattern of drug responses is most similar for agents interfering with DNA replication (idarubicin, cytarabine) and tubulin polymerization (vincristine). These differ however significantly from asparaginase, dexamethasone and venetoclax, revealing reciprocal sensitivities. These differential responses to components of the classical induction and consolidation protocol reinforce the idea of independent drug action where complementary activities increase the likelihood of active combinations for most patients^37,38^.

Leukemia cell survival in co-culture on stroma was a result of the balance between proliferation and cell death, with notable heterogeneity across patients. Proliferation was evident if the cell number in DMSO controls increase over the assay time course. We calculated a proliferation index as the ratio of viable leukemia cells at the endpoint (*t*_72_) relative to the timepoint of drug application (*t*_0_, **Extended Data Fig. 1**). As expected for drugs targeting cell cycle and DNA replication, the proliferation index inversely correlated with logAUC values. Drugs acting on signaling cascades, metabolism or apoptosis were proliferation-independent (**Fig. 3 i/j**). Thus, DRP captures inter-patient diversity in proliferation beyond molecular subtype classification.

### Subtype-specific drug sensitivities delineate actionable vulnerabilities

We next contextualized drug sensitivity within the genomic landscape of ALL^4,5,41^ (**Fig. 4a**). Our cohorts were enriched for high-risk genetic subtypes, including KMT2A rearranged (*n*=33), BCR::ABL1 (*n*=11), TCF::HLF3 (*n*=8), relapsed TCF3::PBX1 (*n*=9), hypodiploidy (n=7), PICALM::MLLT10 and iAMP21 (*n*=3). As expected, BCR::ABL1 and TCF3::PBX1 samples exhibited marked sensitivity to dasatinib and ponatinib (p<0.0001 and p=0.0045, vs. other BCP-ALL), reflecting dependency on ABL/SRC kinase signaling (**Fig. 4b**)^23,42,43^. Relapsed TCF3::PBX1 samples, characterized by higher proliferation rates, also responded to agents targeting mitosis and DNA synthesis (vincristine, barasertib, cytarabine, gemcitabine, mitoxantrone, idarubicin, **Extended Data Fig. 5**). In comparison, TCF3::HLF leukemias were highly venetoclax-sensitive (p=0.0025), consistent with BCL2 transcriptional regulation by the fusion protein (**Fig. 4c**)^44^. Similar dependencies were seen for PICALM::MLLT10^45^, hypodiploidy and KMT2A-rearranged subtypes relative to other ALL (p=0.0096 and p=0.0075 and p=0.018 respectively). KMT2A-rearranged ALL showed age-related differences in venetoclax sensitivity consistent with findings in an independent, age-inclusive cohort^46^. Finally, we also recapitulate sensitivities of early T-cell precursor (ETP) ALL to BCL2 inhibition^47^. (**Extended Data Fig. 5**).

**Fig 4.**
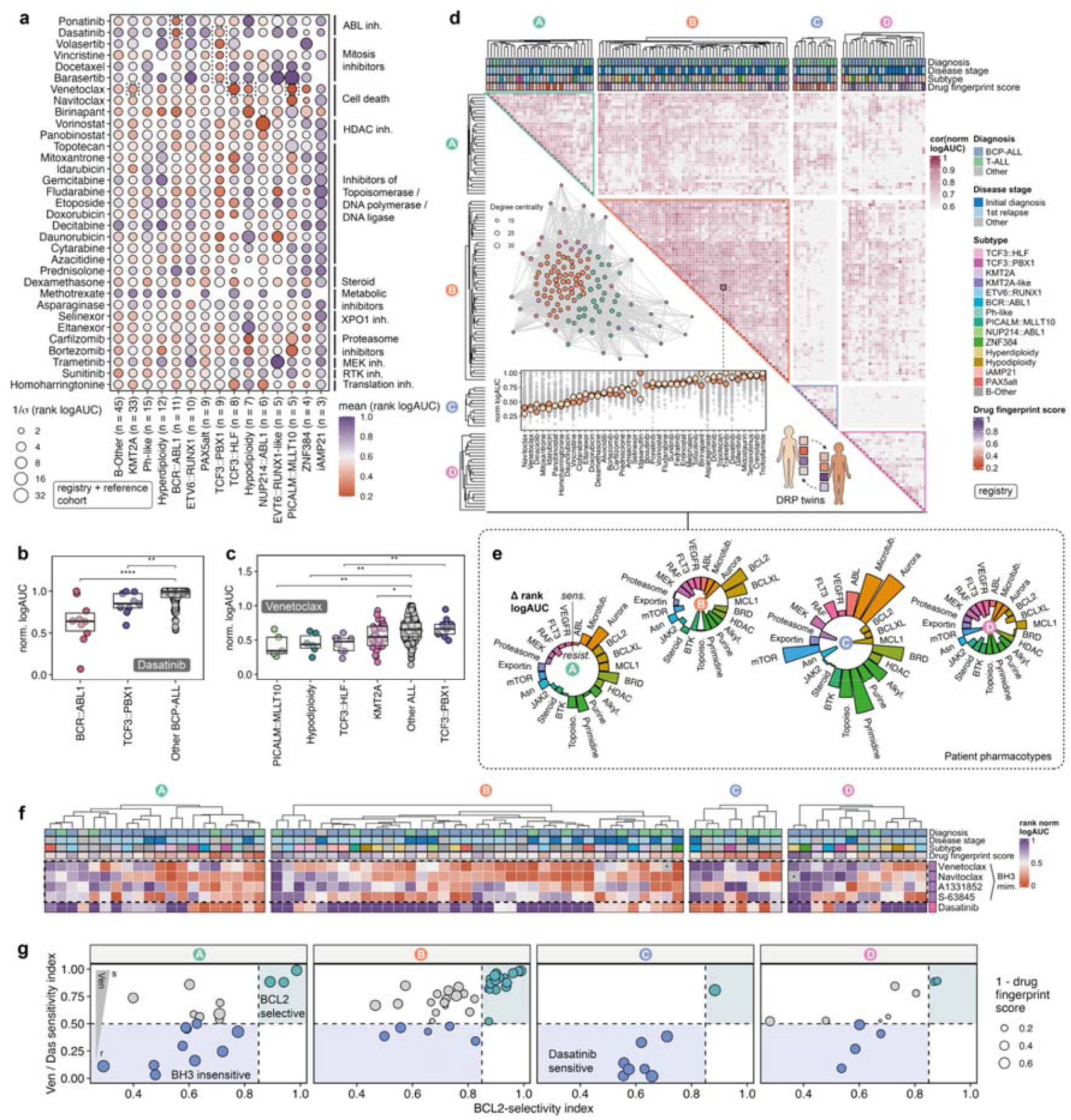
Characterization of subtype-dependent drug sensitivities and identification of DRP twins. **a** Heatmap of averaged ranked logAUC per molecular subgroup. Intra-subtype consistency in drug response is encoded by the size of the circles. **b** Dasatinib responses in selected subtypes (Wilcoxon rank-sum test). **c** Responses to venetoclax in hypodiploidy, PICALM::MLLT10, KMT2A and TCF3 rearranged leukemias. **d** Patient similarity map based on pairwise correlation of drug profiles. The similarity graph connects patients with covarying drug fingerprints identifying phenotypic DRP twins. *K*-means clustering detects four main patient pharmacotypes. **e** Characterization of pharmacotypes by aggregating differential drug ranks across functional classes. **f** BH3-mimetic drug profiling juxtaposing responses to BCL2, BCL-xL and MCL1 inhibitors and in relation to dasatinib. **g** BCL2 sensitivity vs. selectivity index calculated as fractional drug ranks (Eqs. 7 and 8) over the four pharmacotypes.

### Drug fingerprinting identifies functional DRP ‘twins’ independent of genetic classification

To uncover genotype-independent similarities, we computed pairwise correlations between drug fingerprints. Highly correlated fingerprints identified phenotypic ‘DRP twins’ irrespective genetic background (**Fig. 4d**). *K*-means clustering functionally aggregated patients with covarying fingerprints, yielding four main patient pharmacotypes. Patients in cluster A were generally sensitive to conventional chemotherapeutics and partially responded to BCL2/BCL-xL inhibitors. Cluster B showed highest dependence on BCL2 family proteins. Patients in cluster C were susceptible to inhibitors of ABL1, aurora kinase, mTOR and spindle formation, yet they resisted BCL2/BCL-xL inhibition. Cluster D encompassed broadly drug-resistant cases. Moreover, analysis of four BH3 mimetics, namely venetoclax (BCL2 selective inhibitor), S-63845 (MCL1), A1331852 (BCL-xL) and navitoclax (dual BCL2/BCL-xL) revealed subtle heterogeneity in anti-apoptotic dependencies (**Fig. 4f/g**)^47^. Collectively, BH3 mimetics were inversely correlated with tyrosine kinase inhibitors (cluster B vs. cluster C). While cluster B was predominantly composed of BCP-ALL, dasatinib sensitive T-ALL were particularly enriched in cluster C. These observations are consistent with recent work describing LCK activation with concomitant BCL2 downregulation^42^ and linking dasatinib sensitivity to poor prognosis in newly diagnosed T-ALL^23^.

### Functional profiling informs precision treatment decisions in refractory ALL

To evaluate the clinical impact of functional drug screening, we analysed treatments implemented after reporting DRP results to the treating physician. Among all registry and reference patients, 63 patients received at least one non-conventional DRP-tested drug (**Fig. 1, Fig. 5** and **Extended Data Fig. 5**). Most targeted treatments included venetoclax (*n*=37 patients) combined with varying backbones of chemotherapy. A series of refractory T-ALL (*n*=9) received the BCL2 inhibitor in combinations with bortezomib. Patients with measurable clinical responses to venetoclax-based treatment elements exhibited higher *ex vivo* sensitivity than those with progressive disease (p < 0.008, **Fig. 5a/b**). Of 21 patients ranking in the first tertile (≤P33) of *ex vivo* venetoclax sensitivity in the full cohort, 17 (68%) had an objective response (68%). In total, 25 of 37 (68%) venetoclax treated patients were bridged to HSCT or CAR-T cell immunotherapy with 16 patients alive given a median follow-up of 24.3 months after DRP (IQR: 16.2-26.1 months, **Fig. 5c**). Best responders to venetoclax (≤P33, logEC50 <100 nM) showed superior 1-year event-free survival (log-rank p=0.007, 0.57 [95% CI, 0.39-0.85] vs. 0.25 [95% CI, 0.11-0.58]). **Fig. 5d/e**). Illustrative patients included a frontline ETP T-ALL with a PICALM::MLLT10 fusion (patient T-1218) and a relapsed pre-T ALL with TP53/NOTCH1/NRAS mutations (patient T-1212), both remaining in molecular remission 20 and 25 months after DRP respectively (**Fig. 5f/g**). Among 17 TKi-treated patients following drug screening, 13 (76%) had objective responses allowing consolidation with cellular therapy in 11 patients (65%, **Fig. 5h-j**). Besides confirming sensitivities in BCR::ABL1 and NUP214::ABL1 subtypes, DRP further informed the use of ponatinib in a rare case featuring a CNTRL::ABL fusion gene (patient B-0961) as well as in several primary resistant or relapsed T-ALLs without prior knowledge of molecular drivers.

**Fig 5.**
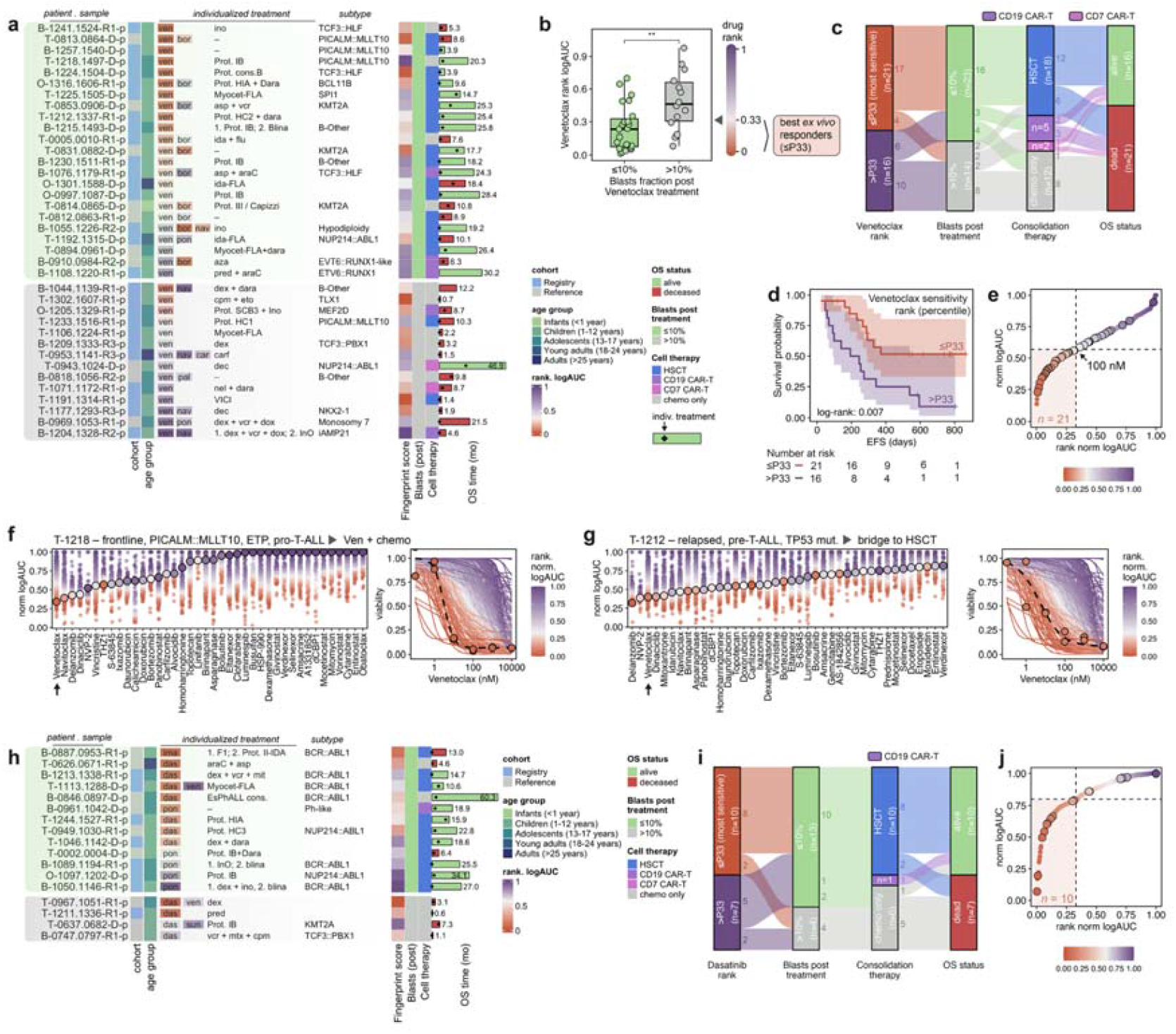
DRP informs individualized therapy selection. **a** Series of relapsed/refractory patients receiving a venetoclax augmented treatment regimen grouped by objective response. DRP informed therapy elements are shown alongside chemotherapy backbones. **b** Patients in with objective responses (≤10% blasts) are significantly more sensitive to venetoclax *ex vivo*. **c** Alluvial plot showing proportions of patients bridged to cell therapy. **d** Event-free survival stratified by *ex vivo* venetoclax sensitivity. **e** Ranked logAUC highlighting patients that received venetoclax and score in the first tercile of the drug distribution. **f** Exemplary case of a ETP patient (T-1218) with exceptional sensitivity to venetoclax and treated on a venetoclax augmented chemotherapy regimen in firstline. **g** Relapsed patient (T-1212) with a TP53 mutation receiving venetoclax as a bridge to HSCT. **h** Patient series treated with dasatinib, ponatinib or imatinib. **i** Alluvial plot with fraction *ex vivo* responders and successful bridging to cell therapy. **j** Ranked logAUC of dasatinib highlighting patient with TKi-based interventions and scoring in the first tercile of the Dasatinib distribution. Drug abbreviations: ven = venetoclax, das = dasatinib, pon = ponatinib, ima = imatinib, vcr = vincristine, araC = cytarabine, dec = decitabine, aza = azacitidine, nel = nelarabine, mit = mitoxantrone, dox = doxorubicin, eto = etoposide, blina = blinatumomab, dara = daratumumab, ino = inotuzumab-ozogamicin.

Collectively, these data demonstrate that functional drug profiling can meaningfully inform individualized therapy, even in heavily pretreated and biologically aggressive ALL. Clinical benefit including reduction of toxicity in frontline and palliative settings, was observed across multiple drug classes, most notably with BCL2 inhibitors and ABL-class TKis but also few examples of MEK and XPO1 inhibitors. These findings validate the clinical utility of DRP in the salvage setting and highlight its translational potential to bridge patients to curative-intent interventions such as transplantation or CAR-T cell therapy. Complementing standard-of-care diagnostics, DRP provides a functional layer for real-time, data-driven therapy selection in relapsed and refractory childhood ALL.

## Discussion

Here, we characterized the drug response landscape of relapsed and refractory pediatric acute lymphoblastic leukemia (ALL). DRP mapped functional dependencies across pediatric B- and T-lineage ALL, revealing new therapeutic opportunities. By profiling 340 patients with r/r ALL across 17 European countries, we demonstrated that centralized, academic DRP is feasible, reproducible and clinically actionable within a federated international network.

Drug fingerprints mirrored treatment response dynamics observed in the clinic. *Ex vivo* resistance to glucocorticoids has long been associated with poor response and early relapse, underscoring that intrinsic and acquired resistance drive adverse outcomes in high-risk ALL^39^. In our cohort, patients with poor prednisone pre-phase response exhibited resistance not only to glucocorticoids but also to drugs targeting apoptotic and stress-response pathways. Likewise, persistent MRD during first-line induction correlated with reduced sensitivity to steroids, anthracyclines, and asparaginase, extending prior evidence linking these sensitivities to MRD kinetics in treatment-refractory disease^23^. These findings reinforce that chemotherapy efficacy is shaped by composite, patient-specific vulnerabilities rather than uniform sensitivity patterns. In line with recent modelling of combination therapy effects, our data suggest that durable efficacy of multidrug regimens arises primarily through additive and complementary mechanisms rather than universal synergy^38,48^. Functional profiling deconvolutes the effects of single drugs and enables rational augmentation of standard backbones with individualized strategies.

By unsupervised clustering of drug response fingerprints, we identified four major pharmacotypes. About half of the samples demonstrated sensitivity to BH3 mimetics, consistent with associations of BCL2 dependency and immature leukemic states in ALL and acute myeloid leukemia (AML)^47,49–51^. Dual inhibition of BCL2 and BCL-xL by combination of venetoclax and navitoclax achieved encouraging responses in r/r ALL^9^. While the navitoclax program has been discontinued, next-generation agents such as DT2216^52^, LP-118^53^ and mirzotamab clezutoclax^54^ are in clinical development and may soon be available for precision trials. Network analysis of single-cell transcriptomes identified BCL2 activity in association with earlier pre-proB lineage states in newly diagnosed ALL^55^. We showed that TCF3::HLF, PICALM::MLLT10, KMT2A-rearranged and hypodiploid ALL were most sensitive to venetoclax, yet exceptional responders are observed across molecular subtypes. This is consistent with increased levels of BCL2 in these subtypes^44,45,56,57^ and likely reflects subclonal diversity in lineage maturation and apoptotic priming^50,58^. DRP reliably captured BCL2-family dependencies suggesting that high-content, small-molecule perturbation profiling can serve as a scalable complement to BH3 profiling^59^, while providing information across a wide range of drugs. Conversely, a distinct subset of T-ALL shared fingerprints characterized by sensitivity to dasatinib, aurora kinase, and mTOR inhibitors but marked BH3 mimetic resistance, in agreement with previously reported dasatinib-responsive T-ALL^19,23^. Notably, we documented successful interventions in patients treated with dasatinib based combinations following DRP, illustrating the translational relevance of our approach.

Recent work has revealed substantial biological heterogeneity within established molecular subtypes of ALL^58^, including the presence of HSPC-like subpopulations linked to clinical treatment resistance and poor outcome^50,55^. Similarly, single-cell RNA sequencing detected a rare stem-like and drug-resistant subpopulation already present at diagnosis that expanded at relapse^60^. These insights underscore the need for functional precision oncology linking pharmacotypes with molecular and single-cell layers to resolve subclonal and lineage-associated diversity. DRP assays can couple viability readouts with multiplexed fluorescent and label-free imaging to characterize morphological and subcellular responses^27,61,62^ and inform combination design^63–65^. Furthermore, artificial intelligence based analysis of complex genomic data is improving disease classification and risk stratification in hematologic malignancies, including ALL and myelodysplastic syndromes (MDS)^66–68^. The next frontier for functional precision oncology is the implementation of real-time, data-driven decision making in tumor boards on the basis of integrated genomic and functional datasets. To foster collaboration across borders and legislations, federated approaches are promising^69,70^.

Based on our harmonized laboratory workflows and standardized drug scoring, clinical trials can be developed to benchmark the impact of DRP. Windows for evaluating DRP-guided interventions in ALL include MRD persistence during frontline or relapse therapy, such after blinatumomab treatment, and later refractory disease setting. Our registry experience matches with other international precision oncology initiatives^17^, indicating that earlier integration of functional profiling may broaden therapeutic benefit before resistant subclones emerge. The primary clinical objective is to improve depth of remission for consolidation with cellular therapies such as HSCT or CAR-T. Furthermore, DRP-selected therapy elements may reduce toxicity also in palliative settings.

In conclusion, our data support the feasibility and actionability of real-time drug profiling for clinical decision-making in childhood leukemia. Our results illustrate the potential of DRP-guided interventions with objective responses across multiple targeted drug classes, reducing leukemia load and bridging to cellular therapies. This constitutes the basis for introducing DRP in adaptive, matrix-based studies to assist selection of individualized treatment for patients with r/r ALL.

## Data Availability

Data produced in the present study are available upon reasonable request to the authors.

## Author contributions

F.D.S., A.L., B.C.B. and J.P.B. designed the study. F.D.S, A.L., N.S. setup case report forms. A.L., J.A., N.S., N. B., C.E., A.Z., B.T.A.S., A.A., A.R., S.E., E.J., B.D.M., P.S., P.A., L.Sr., B.B. A.B., J.M.B., M.M., F.C., M.A., and J.H. registered and treated patients. A.L., N.S., L.S and S.S. collected patient CRF and curated clinical data. L.S., A.A., S.G. and D.M optimized DRP wet-lab workflows, performed co-clinical DRP assays and prepared reports. N.S., N.B. and J.P.B led clinical case discussions. A.K., I.V., G.G., F.K. and P.Ho implemented the image analysis pipeline with BIAS. F.D.S. and N.B. built the DRP analytical framework. F.D.S analyzed the data and created the figures. K.S., P.R., F.L., F.dB., M.S., A.E.K., M.S., H.C., A.B., A.vS. and G.C. contributed patients and advice for the study. J.P.B and B.C.B supervised the study. F.D.S. and J.P.B. wrote the manuscript. All authors reviewed the manuscript.

## Acknowledgments

The authors thank all treating physicians of the “*ALL resistant disease consortium”* for referring patients to DRP and insightful case discussions. Funding from the University of Zurich (ZPOC, UMZH), individualized Pediatric Cure (iPC, Horizon Europe, grant no. 826121), IntReALL2020 (Horizon Europe, MISS-2022-Cancer-01, grant no. 101104582), The LOOP Zurich (INTeRCePT), Forschungszentrum für das Kind (FZK, Heidi Ras) of the University Children’s Hospital Zurich as well as financial support from various foundations are kindly acknowledged. The authors would like to thank all patients and their families.

## ALL resistant disease consortium

Brigitte Klug Albertsen^31^, Bodil E. Thorhauge Als-Nielsen^7^, Julia Alten^3^, Philip Ancliff^13^, Marie Angoso^32^, Marc Ansari^21^, Andishe Attarbaschi^8^, Adriana Balduzzi^16,17^, Frederic Baleydier^21^, André Baruchel^29^, Nicole Bodmer^1^, Judith M. Boer^18^, Beat C. Bornhauser^1^, Jean-Pierre Bourquin^1^, Nimrod Buchbinder^33^, Barbara Buldini^15^, Gunnar Cario^3^, Hélène Cavé^29^, Francesco Ceppi^20^, Roman Crazzolara^34^, Laurence Dedeken^35^, Francesca DelBufalo^23^, Ximo Duarte^36^, Cornelia Eckert^2^, Sarah Elitzur^9^, Jonas Grauhan^2^, Jeanette Greiner^37^, Bernd Gruhn^38^, Jörg Halter^22^, Birte Heller^3^, Shai Izraeli^10^, Elad Jacoby^10^, Axel Karow^39^, Andreas E. Kulozik^26,27^, Tim Lammens^11^, Thierry Leblanc^29^, Lennart Lenk^3^, Andrej Lissat^2^, Franco Locatelli^23,24^, Roland Meisel^40^, Monika Mielcarek^19^, Barbara De Moerloose^11^, Brigitte Nelken^41^, Anna Nilsson^12^, David O’Connor^13^, Marlene Pasquet^42^, Tomaz Prelog^30^, Annika Rademacher^2^, Alexandra Reekmans^11^, Raffaele Renella^20^, Carmelo Rizzari^17^, Pierre Rohrlich^28^, Nastassja Scheidegger^1^, Paul-Gerhardt Schlegel^43^, Kjeld Schmiegelow^7^, Martin Schrappe^3^, Heidi Segers^44^, Saskia Sonnenberg^3^, Lucie Sramkova^14^, Arend von Stackelberg^2^, Martin Stanulla^25^, Fabio D. Steffen^1^, Petter Svenberg^12^, Anna Valaine^45^, Nicolas von der Weid^22^, Koray Yalcin^46^, Aida Zeckanovic^1^

^*31*^*Aarhus University Hospital, Denmark*, ^*32*^*CHU Bordeaux, France*, ^*33*^*CHU Rouen, France*, ^*34*^*Universitätsklinikum Innsbruck, Austria*, ^*35*^*Hôpital Universitaire de Bruxelles, Belgium*, ^*36*^*University Medical Centre Ljubljana, Slovenia*, ^*37*^*Kantonsspital Aarau, Switzerland*, ^*38*^*Universitätsklinikum Jena, Germany*, ^*39*^*Universitätsklinikum Erlangen, Germany*, ^*40*^*Universitätsklinikum Düsseldorf, Germany*, ^*41*^*CHU Lille, France*, ^*42*^*CHU Toulouse, France*, ^*43*^*Universitätsklinikum Würzburg, Germany*, ^*44*^*UZ Leuven, Belgium*, ^*45*^*Children’s Clinical University Hospital Riga, Latvia*, ^*46*^*Acibadem Altunizade Hospital, Turkey*

## Extended Data Figures

**Extended Data Figure 1.**
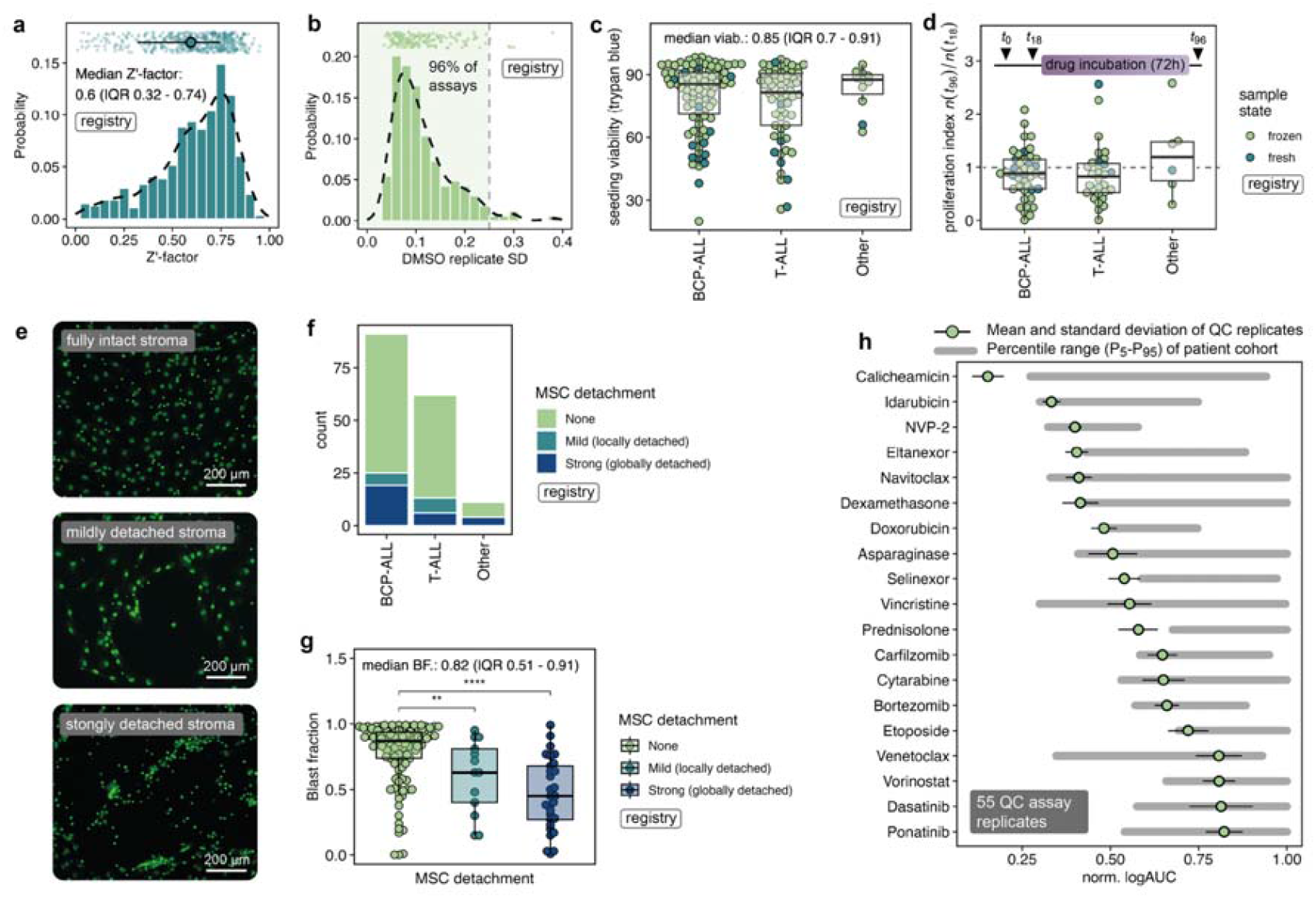
Sample viability and assay quality control. **a** Distribution of plate-wise Z’-factors quantifying assay reliability by the separation of positive (idarubicin) and negative (DMSO) controls. **b** Distribution of the standard deviation of DMSO cell counts averaged across all plates for each assay. 96% of all assays in the registry cohort had an average DMSO standard deviation <0.25. **c** Cell viability measured by trypan blue staining before seeding the leukemia cells on the mesenchymal stromal cell (MSC) monolayer. **d** Proliferation index calculated as the ratio of viable cells counted by CyQuant prior to drug application (*t*_0_) and after 72 h of drug incubation (*t*_72_). **e** Representative images of MSC monolayers. **f** Degree of MSC detachment in untreated ALL wells compared to MSC only controls. **g** Dependence of MSC detachment on blast fraction. Samples with lower blasts percentages show significantly higher degrees of detachment (Wilcoxon test, p=0.008 and p<0.0001). **h** DRP reproducibility with a PDX control sample showing mean±σ of 55 assay replicates. The gray interval indicates the spread of the combined registry and reference cohort (quantiles 0.05-0.95).

**Extended Data Figure 2.**
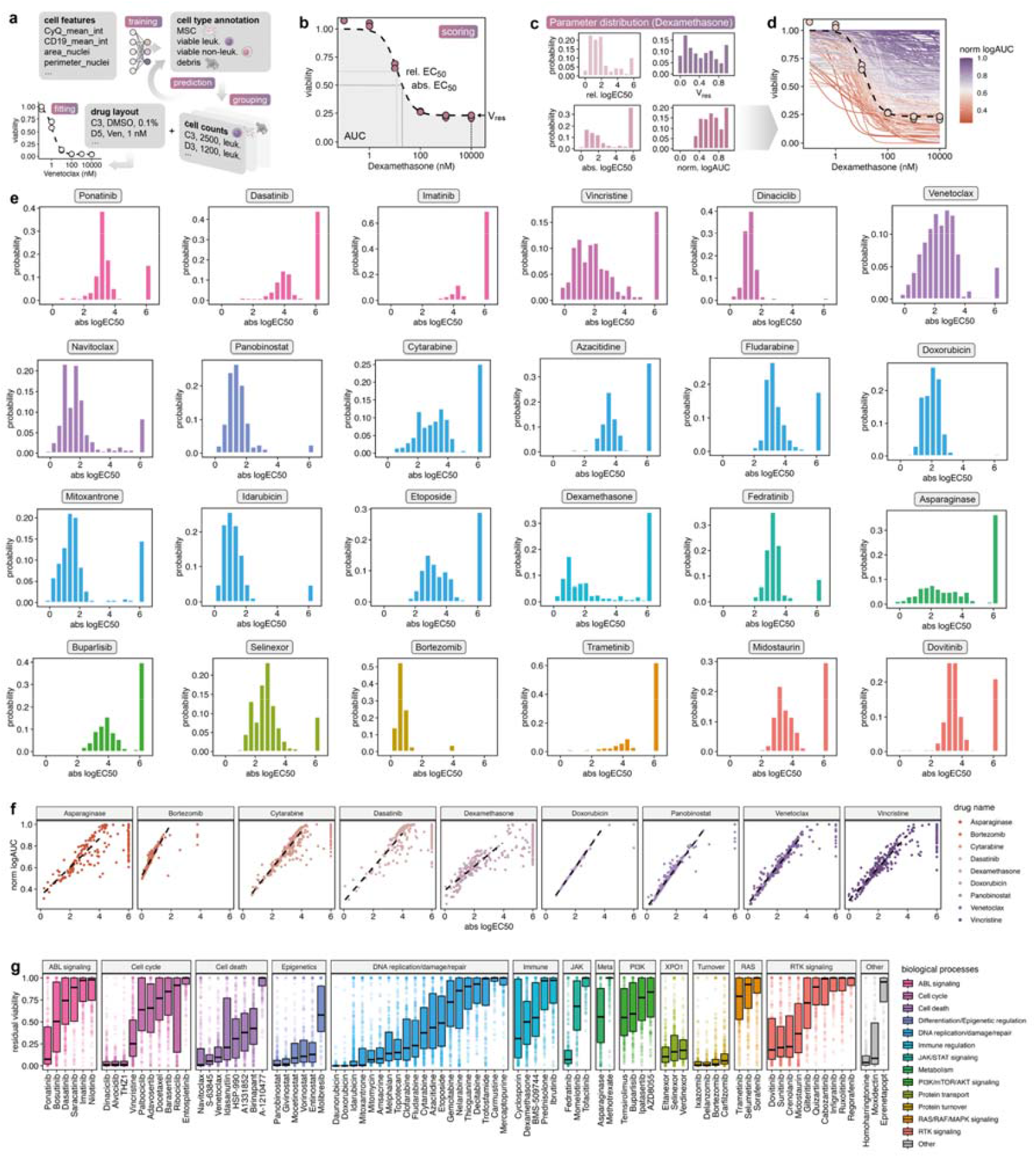
Scoring drug activity using image-based DRP. **a** Schematic of computational workflow for feature extraction, cell type annotation and drug response calculation. **b** Parameters derived from drug response fits and used for scoring. The residual viability *V*_res_ quantifies the remaining cell fraction at the highest measured concentration relative to vehicle controls (DMSO). **c** Parameter distribution of relative and absolute logEC_50_ (details in Online Methods), *V*_res_ and normalized logAUC, exemplified for dexamethasone. **d** Drug responses of dexamethasone color-coded by the normalized logAUC. Each line corresponds to a patient sample. **e** Histograms of absolute logEC50 distributions for selected drugs of relevant biological processes in ALL. While many cytotoxic drugs (e.g. anthracyclines, proteasome, BCL2, HDAC inhibitors) showed a monomodal, near-Gaussian activity profile, the distributions of many cytostatic and targeted compounds spanned over several log units (e.g. mitosis inhibitors) or exhibit significant proportions of non-activity (e.g. MEK and (receptor) tyrosine kinase inhibitors). **f** Correlation of absolute logEC50 and normalized logAUC parameters across the measured concentration range. **g** Distribution of residual cell viability at the maximal drug concentration.

**Extended Data Figure 3.**
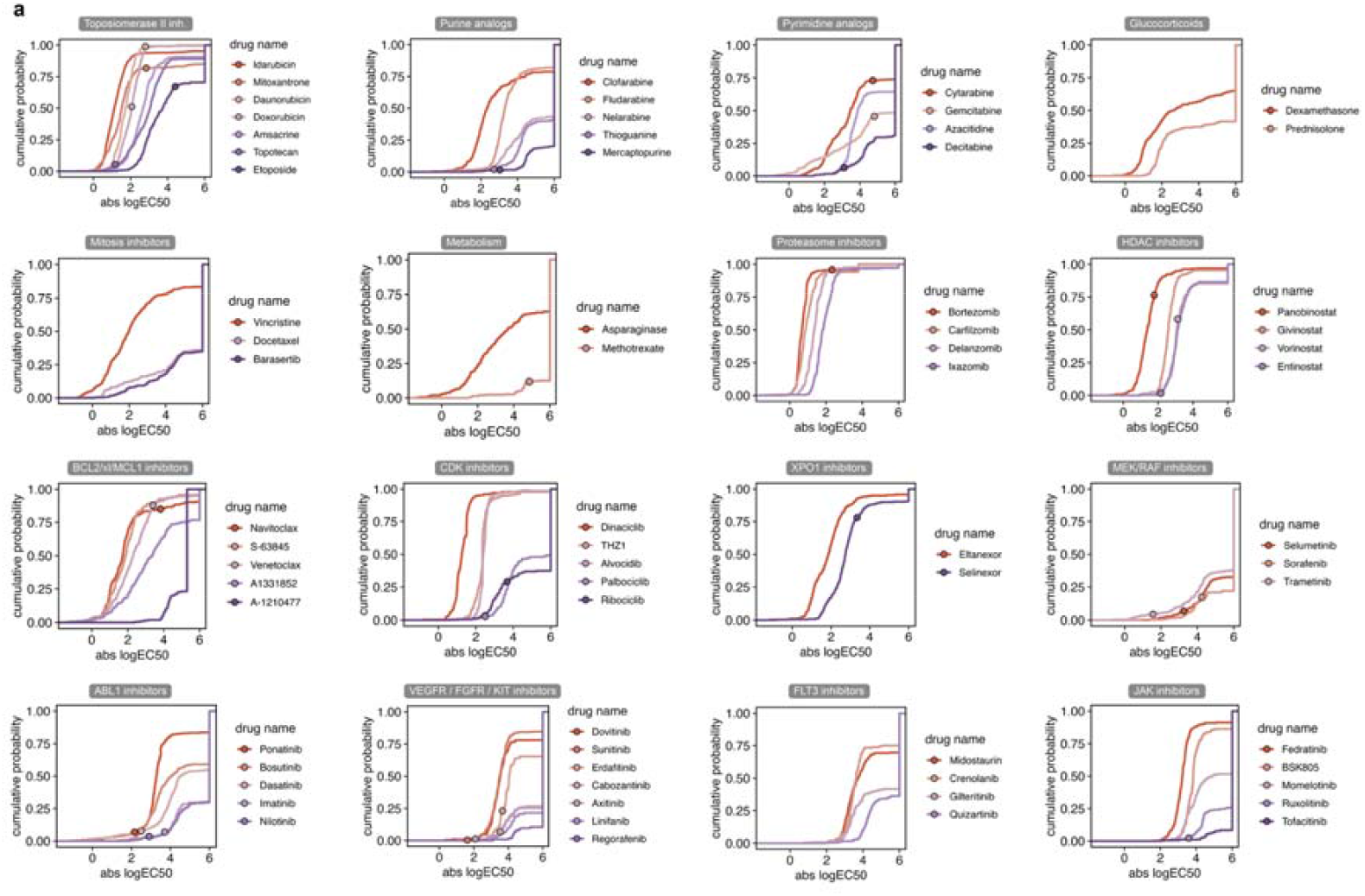
Cumulative probability distributions of absolute logEC50 indicating different activity levels of the drugs in the library. Circles denote the maximum concentration *C*_p_,_max_ where available from literature^36^. The y-axis indicates the fraction of patient samples below any given absolute logEC50. The proportion of patients completely resistant to a drug (abs. logEC50 = 6) varies considerably across and within drug classes.

**Extended Data Figure 4.**
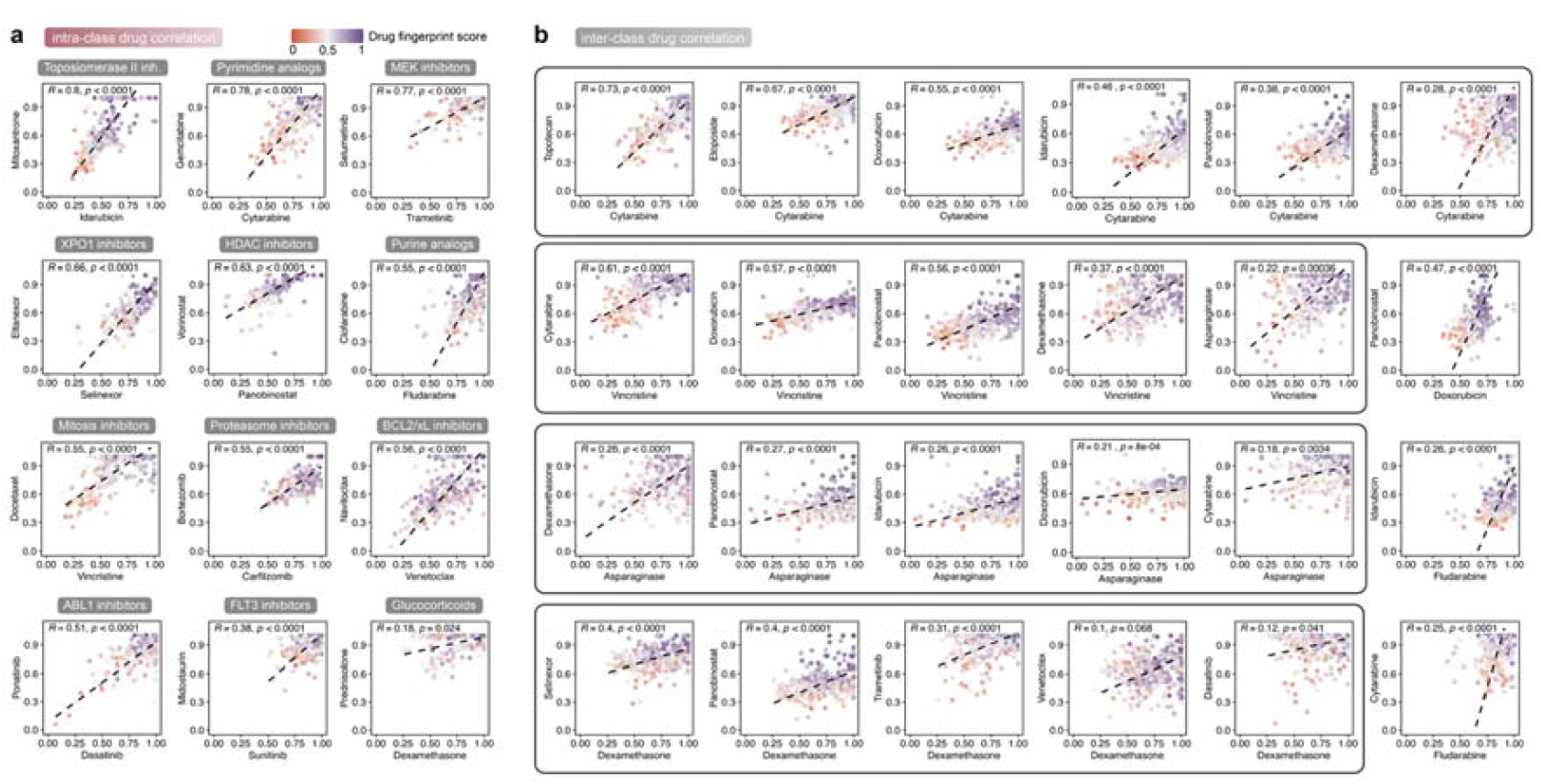
Spearman rank correlation of drugs in the same and disparate functional classes. **a** Intra-class drug correlations corroborate stability of drug responses (intra-class effects). **b** Inter-class correlations indicate orthogonal drug action with potential for drug additivity as suggested by established combinations in induction, consolidation and high-risk blocks of clinical trial protocols.

**Extended Data Figure 5.**
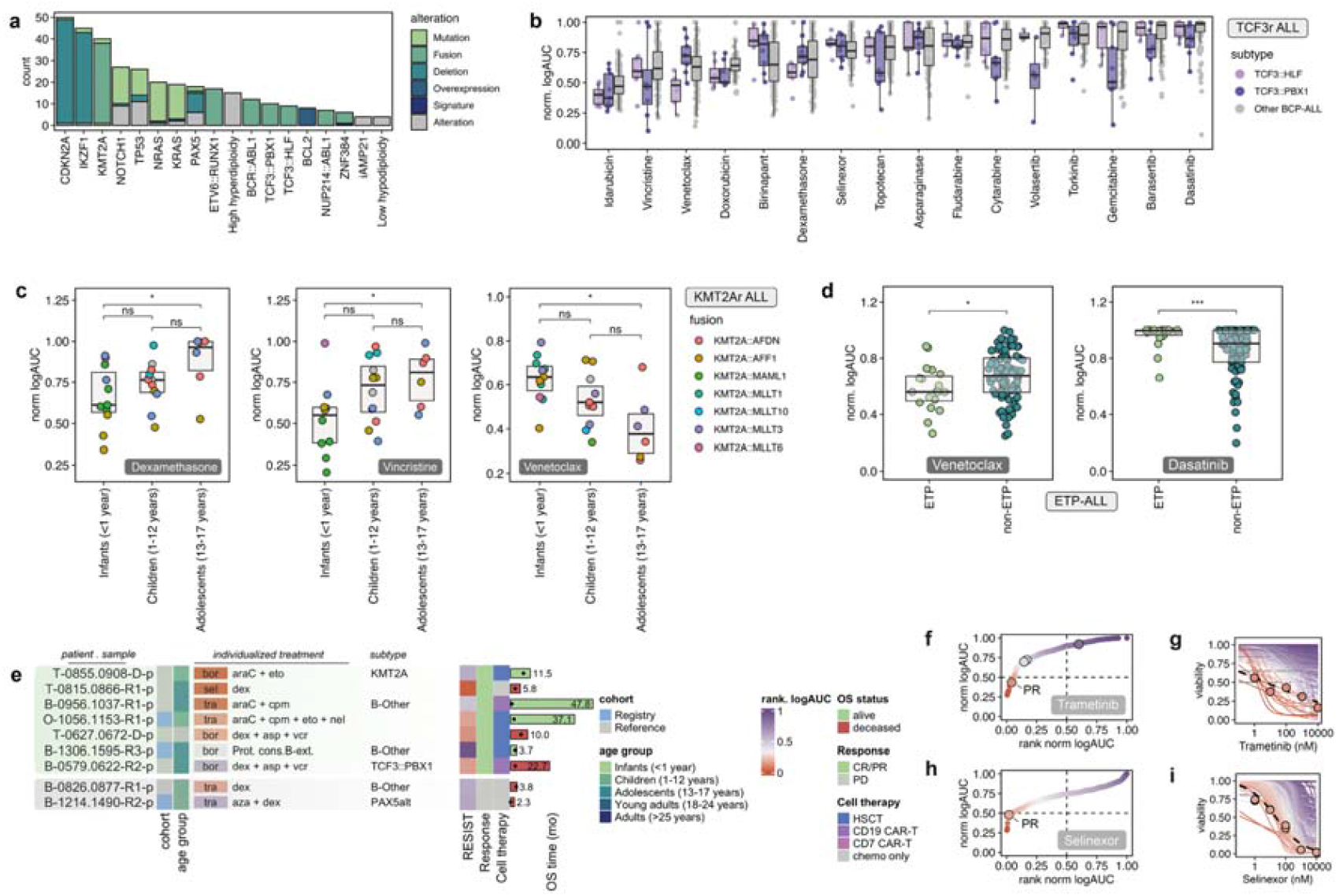
Functional characterization of selected molecular subtypes in pediatric ALL. **a** Frequencies of recurrent molecular alterations (SNVs, fusions, deletions, overexpression) as reported by the treating physician. **b** DRP of differentially active drugs in TCF3 rearranged leukemia. **c** Age-dependence of KMT2A rearranged leukemias for dexamethasone, vincristine and venetoclax. **d** Differential sensitivity to venetoclax and dasatinib in ETP and non-ETP ALL. Group comparisons with Student’s t-test. **e** Patients with targeted treatments following DRP, containing either selinexor (sel), trametinib (tra) or bortezomib (bor).

## Online Methods

### Patient cohort

Patients were enrolled in the drug response profiling (DRP) registry (*NCT06550102*) or profiled as part of add-on projects to clinical trials (reference cohort). Informed consent was given in accordance with the Declaration of Helsinki and the ethics commission of the Canton Zurich, Switzerland (KEK-2021-02189 and KEK-2014-0383). Patients in both cohorts have been treated according to AIOEP-BFM ALL 2009/2017, NOPHO ALL2008, ALLTogether, ALLIC 2009, EsPhALL 2017 first-line studies or on IntReALL SR/HR 2010, ALL-REZ BFM 2002 and UKALL 2011 relapse protocols. Treatment decisions including individualized, DRP-guided precision therapy was at the sole discretion of the referring physician. Drug screening was prioritized by clinical urgency, intent-to-treat and risk stratification as defined by AIEOP-BFM and IntReALL clinical trials. Patient demographic data (sex, age) and basic disease characteristics were collected from the treating physician including current disease stage (initial diagnosis, first, second and high-order relapse), immunophenotype as BCP-ALL, T-ALL/T-LBL, mixed-lineage acute leukemia (MPAL) or acute undifferentiated leukemia (AUL), maturation degree according to EGIL classification as well as ETP status for T-ALL. Genetic subtypes and molecular lesions were annotated if available from the referring center at the time of DRP (**Table 1, Extended Data Table 1** and **Extended Data Fig. 5a**). Treatment documentation in first-line therapy included (i) prednisone response to the cytoreductive pre-phase at day 8 with an absolute cell count of 1000 blasts/μL in peripheral blood defining poor response, (ii) flow-cytometry MRD at day 15 of therapy induction, with a cutoff at ≥10% blasts in bone marrow, and (iii) end-of-induction bone marrow assessment (day 33) by morphology (non-remission defined at ≥5% blasts) and PCR-MRD. Follow-up after DRP including individualized drug treatments, hematopoietic stem-cell transplantation and/or CAR-T therapy with associated response assessments was provided by the treating physician. Event-free survival (EFS) was defined as the time from bone marrow sampling for DRP to either relapse or death from any cause. Likewise, overall survival (OS) was defined as the time from the bone marrow puncture to death. Patients not experiencing the event were right-censored at the time of last follow-up. The survival function was modeled using the Kaplan-Meier estimator (*survival* package in R) and differences in survival distributions were assessed with the log-rank test.

### Cell culture and drug screen

A monolayer of 2500 hTERT-immortalized primary bone marrow mesenchymal stroma cells (MSC) in AIM V medium were plated per well in a 384 multi-well format (black μCLEAR®, Greiner) using a MANTIS liquid dispenser (Formulatrix) or a multichannel pipette. MSC cultures were incubated overnight. Primary human ALL cells were recovered from cryotubes or isolated from fresh bone marrow aspirates or peripheral blood. On average, 10000 leukemia cells were seeded onto the stromal layer and the co-culture was incubated overnight allowing the cells to settle. The compound library with up to 113 drugs including conventional chemotherapeutics and molecularly targeted agents was reconstituted in 10 mM DMSO or H_2_O and stored as stock plates at -80°C (**Extended Data Table 2**). Drugs were added in 5- or 6-point serial dilutions using an HP-D300 Digital dispenser (Tecan) and OT-2 liquid handler (Opentrons) or an ECHO 650 acoustic liquid handler (Beckman Coulter). Most compounds were measured on a logarithmic dilution scale between 0.1 – 10000 nM, encompassing physiological plasma levels^36^. For highly potent drug substances, concentrations were optimized separately (calicheamicine: 0.001 – 2.5 nM, proteasome inhibitors: 0.02 – 62.5 nM). Dispending volumes were adjusted to keep DMSO concentrations ≤0.125%. Cells were exposed to drugs for 72 h before staining with CyQuant® reagent (ThermoFisher, dye diluted 1:600, repressor diluted 1:40 for a 4X master mix) to assess cell viability. Selected samples with low blast fractions and signs of detaching MSCs were additionally stained with allophycocyanine (APC) conjugated antibodies against CD19 (SJ25C1) or CD7 (124-1D1) to identify leukemia cells.

### Automated microscopy and image analysis

After 45 min incubation with CyQuant, cells were imaged on an Operetta CLS (Revvity, previous PerkinElmer) or an ImageXpress Micro (Molecular devices) high-content fluorescence microscope in widefield at 10x magnification with excitation and emission bandpass filters set at 460-490 nm and 500-550 respectively. Images were analyzed using a machine learning pipeline described previously^71,72^. Cells were segmented and classified as live leukemia, MSC or cellular debris using the Advanced Cell Classifier^71^ or the Biological Image Analysis Software (BIAS, Single-Cell Technologies)^72^. In brief, images were normalized by defining underflow and overflow bins on the summed histogram of all images to account for background and outlier pixel intensities. Cells with areas in a range of 10-3000px were identified using a leukemia segmentation model, based on the nucleAIzer Mask-RCNN^73^ model and implemented in BIAS^72^. Normalized intensities, area, perimeter and texture features were extracted and used to train a supervised classification model in recognizing MSCs and differentiating between viable and dead leukemia cells. Cell type annotations were collected across multiple samples and drug conditions to serve as a ground training set. Predictions were visually inspected and out-of-distribution samples showing cell clustering or pronounced detachment of the stromal layer were retrained on-the-fly. Total cell counts were aggregated per condition and compiled for downstream dose-response analysis.

### Drug response scoring

Drug responses were normalized plate-wise to DMSO-treated negative control wells. The measured dose responses were fitted to a three-parameter model (logEC_50_, *E*_max_, *n*) using the *drc* R package^74^. Extreme outlier which abrogated the curve fits were manually masked from logistic regression but kept as data points in the profiles. Goodness of fit was estimated by the residual square error (RSE) of the measured data points to the regression line, as recently described^75^. The *relative* logEC_50_ was defined as the inflection point of the sigmoidal curve (concentration with half-maximal effect relative to the maximal effect *E*_max_), while the *absolute* logEC_50_ was determined as the concentration where the drug effect is exactly 50%). Relative and absolute logEC_50_ converge with *E*_max_ approaching 0. The absolute logEC_50_ was capped symmetrically at log_10_(*c*_min_) – 2 and log_10_(*c*_max_) + 2. The residual cell viability *V*_res_ was defined as the relative cell fraction at the maximally measured concentration.

We used the area under the logistic response curve as an integrated metric for drug response^76,77^. The logarithmic area-under-the curve (logAUC) was calculated as the analytical definite integral of the four-parameter log-logistic model

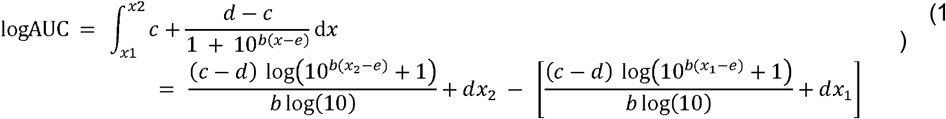

The logAUC was normalized to the tested concentration range for each drug as

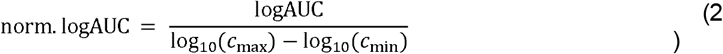

Z-transformations were computed per drug for both absolute logEC_50_ and logAUC as 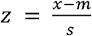 where *μ* and *σ* refer to the mean and standard deviation of the metric *x* across the patient cohort.

To identify exceptional responders to any given compound, a drug-wise ranking score was assigned to each patient based on the cumulative distribution function (CDF)

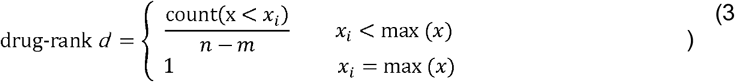

where *n* is the total number of patients in the given cohort and *m* is the number of patients where the drug metric *x* (i.e. the logEC_50_ or the logAUC) is at the maximum. This slight modification of the CDF ensures continuous scaling of the drug rank between >0 (best response) and 1 (non-response). Collectively, the drug ranks generate the drug fingerprint of a patient, represented here as a column vector

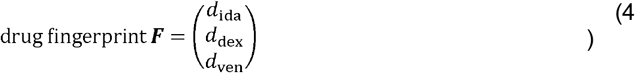

To compute a drug fingerprint score *S*, weights are assigned to each drug based on the variance of the normalized logAUC values in the patient cohort, e.g. for idarubicin

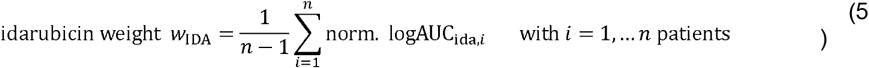

The drug fingerprint score is then calculated as the scalar product of the drug weights and the drug fingerprint

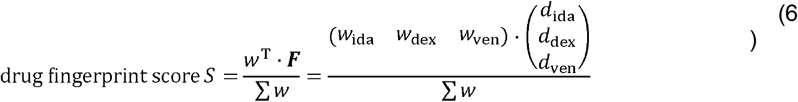

where *w*^T^ is the row vector of drug weights, and ***F*** the drug fingerprint with the measured drug ranks (ida: idarubicin, dex: dexamethasone, ven: venetoclax).

BCL2 inhibitor sensitivity and selectivity indices were compute from fractional drug ranks

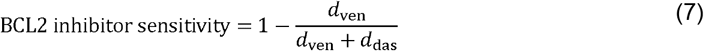

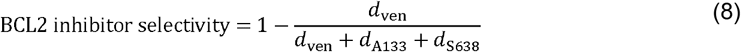

where *d*_DAS_ is the drug rank of tyrosine kinase inhibitor dasatinib, *d*_A133_ the BCL-xL selective inhibitor A1331852 and *d*_S638_ the MCL1 inhibitor S-63845.

### Assay quality criteria

Sample quality was assessed prior to seeding of leukemia cells by measuring baseline cell viability with trypan blue. Samples with <1.5 Mio viable cells at arrival were excluded from DRP. To monitor cell survival and quantify proliferation, untreated cells were imaged with CyQuant before drug addition (timepoint *t*_18_). Integrity of the stroma layer was assessed both visually and quantitatively by determining the degree of surface coverage in wells with leukemia/MSC co-cultures relative to MSC-only conditions. Based on cytoplasmic segmentation of GFP-positive MSCs, detachment of the stroma monolayer was categorized as absent, mild or strong. Consistency in cell counts across wells was surveyed by calculating average standard deviation of the DMSO replicates across all plates. The histogram of plate-averaged DMSO standard deviations displayed a near-Gaussian distribution around a mean of 0.1. We chose a cutoff at σ_DMSO_<0.25 corresponding to the 95^th^ percentile of the distribution. Assays were failed if either the variability in DMSO replicates exceeded this threshold, less than 50 leukemia cells were recovered in DMSO at the endpoint or leukemia cells could not be identified as a result of strong detachment or sample contamination. Drug perturbations were performed in duplicates for each drug concentration condition. Individual drug responses were flagged for deviating technical replicates and masked if curve fits did not converge. The anthracycline idarubicin was present on every drug plate and served as a positive control. *Z*’-factors were calculated plate-wise from the mean and standard deviation of the raw cell counts in DMSO (*μ*_n_ and *σ*_n_) and idarubicin (*μ*_p_ and *σ*_p_) at the highest measured concentration, according to 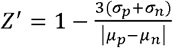. Compounds with exceptionally narrow activity windows (dinaciclib, alvociclib, THZ-1, bortezomib) served as additional controls. Mycoplasma contamination was monitored by monthly PCR testing of MSC cultures, and assays were censored in case of positive testing. To evaluate technical reproducibility of DRP, a T-ALL patient derived xenograft (PDX) sample was repetitively screened with a representative subset of 19 drugs (*n*=55 technical replicates).

### Hierarchical and *K*-means clustering and network analysis

Hierarchical clustering was performed using the unweighted pair group method with arithmetic mean (UPGMA) on the Euclidian distance matrix of the raw normalized logAUC values, ranked logAUC or the pairwise correlation coefficients. Dendrograms were reordered by row and column means and visualized with the *ComplexHeatmap*^78^ bioconductor package. For DRP twin analysis, pairwise Spearman rank correlation coefficients were computed on the patients’ drug fingerprints. A weighted adjacency matrix was calculated by soft-thresholding the similarity matrix using a power of *β*=6 in analogy to weighted gene correlation network analysis (WGCNA)^79^.

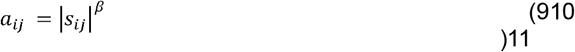

The resulting unsigned network was visualized using the *tidygraph* and *ggraph* packages. Node importance was assessed by degree centrality and edges with weights <0.1 were truncated for clarity. Distinct pharmacotypes were identified by *K*-means clustering. The optimal number of clusters was determined by the plotting the *within cluster sum of squares (WCSS)* as a function of increasing cluster centers.

### Statistical analysis

Statistical significance tests were reported in the main text and corresponding figure legends. For group comparisons two-sided Student’s t-tests or non-parametric Wilcoxon rank-sum tests were used as appropriate depending on data distribution. Significance levels were indicated with asterisks as *p*<0.05 (*), *p*<0.01 (**), *p*<0.001 and *p*<0.0001 (****) with *p*-values corrected for multiple testing using the false discovery rate (FDR) procedure. Boxplots indicate median and interquartile range. Drug and patient correlations were assessed with Spearman rank coefficients and annotated with two-tailed *p*-values. All statistical analyses were conducted in *R* version 4.4.

## Extended Data Tables

**Extended Data Table 1.** Patient characteristics of the combined registry and reference cohorts.

| Patient characteristics | N = 340 |
| --- | --- |
| **Sex** |  |
| Male | 209 (62%) |
| Female | 129 (38%) |
| Not available | 2 |
| **Age group** |  |
| Infants (<1 year) | 18 (6%) |
| Children (1-12 years) | 191 (59%) |
| Adolescents (13-17 years) | 88 (27%) |
| Young adults (18-24 years) | 8 (2%) |
| Adults (>25 years) | 19 (6%) |
| Not available | 16 |
| **Diagnosis** |  |
| BCP-ALL | 189 (56%) |
| T-ALL | 130 (38%) |
| T-LBL | 6 (2%) |
| MPAL | 11 (3%) |
| AUL | 4 (1%) |
| **Disease stage** |  |
| Initial diagnosis | 149 (44%) |
| 1st relapse | 140 (41%) |
| Higher relapse | 51 (15%) |
| **Risk** |  |
| SR (Initial diagnosis) | 25 (7%) |
| MR (Initial diagnosis) | 8 (2%) |
| HR (Initial diagnosis) | 116 (34%) |
| SR (1st relapse) | 16 (5%) |
| HR (1st/higher relapse) | 175 (51%) |
| **Subtype<sup>1</sup>** |  |
| TCF3::HLF | 9 (3%) |
| TCF3::PBX1 | 10 (3%) |
| KMT2A | 35 (10%) |
| ETV6::RUNX1 | 10 (3%) |
| EVT6::RUNX1-like | 5 (1%) |
| BCR::ABL1 | 11 (3%) |
| Ph-like | 15 (4%) |
| PICALM::MLLT10 | 5 (1%) |
| NUP214::ABL1 | 6 (2%) |
| ZNF384 | 4 (1%) |
| Hyperdiploidy | 13 (4%) |
| Hypodiploidy | 7 (2%) |
| iAMP21 | 4 (1%) |
| MEF2D | 3 (1%) |
| PAX5alt | 10 (3%) |
| B-Other | 47 (14%) |
| Unspecified BCP-ALL | 21 (6%) |
| Unspecified T-ALL | 109 (32%) |
| Unspecified MPAL/AUL/T-LBL | 16 (5%) |
| **ETP status** |  |
| non-ETP | 107 |
| ETP | 23 |

<sup>†</sup> As reported by the treating physician.

**Extended Data Table 2.**
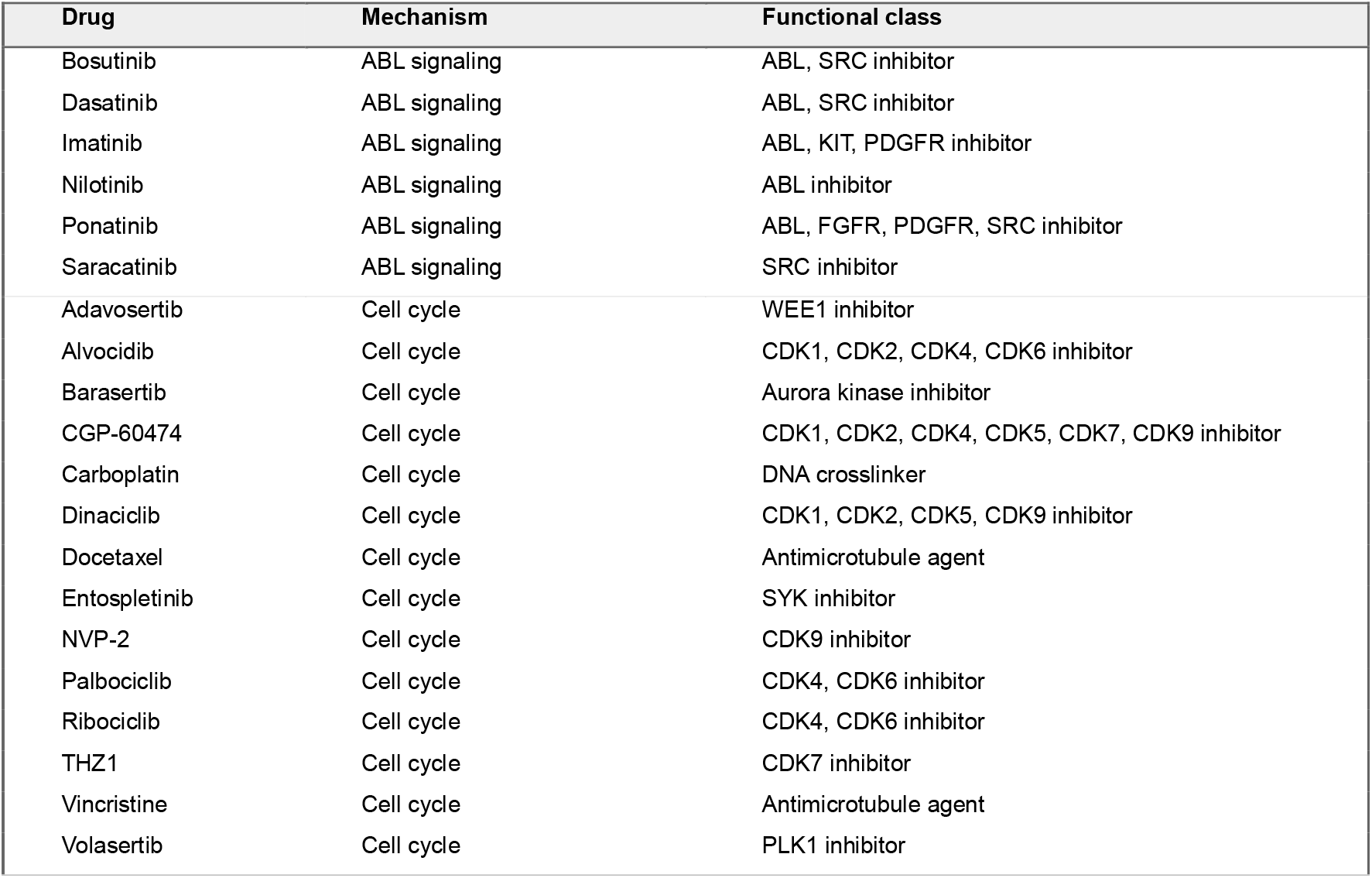
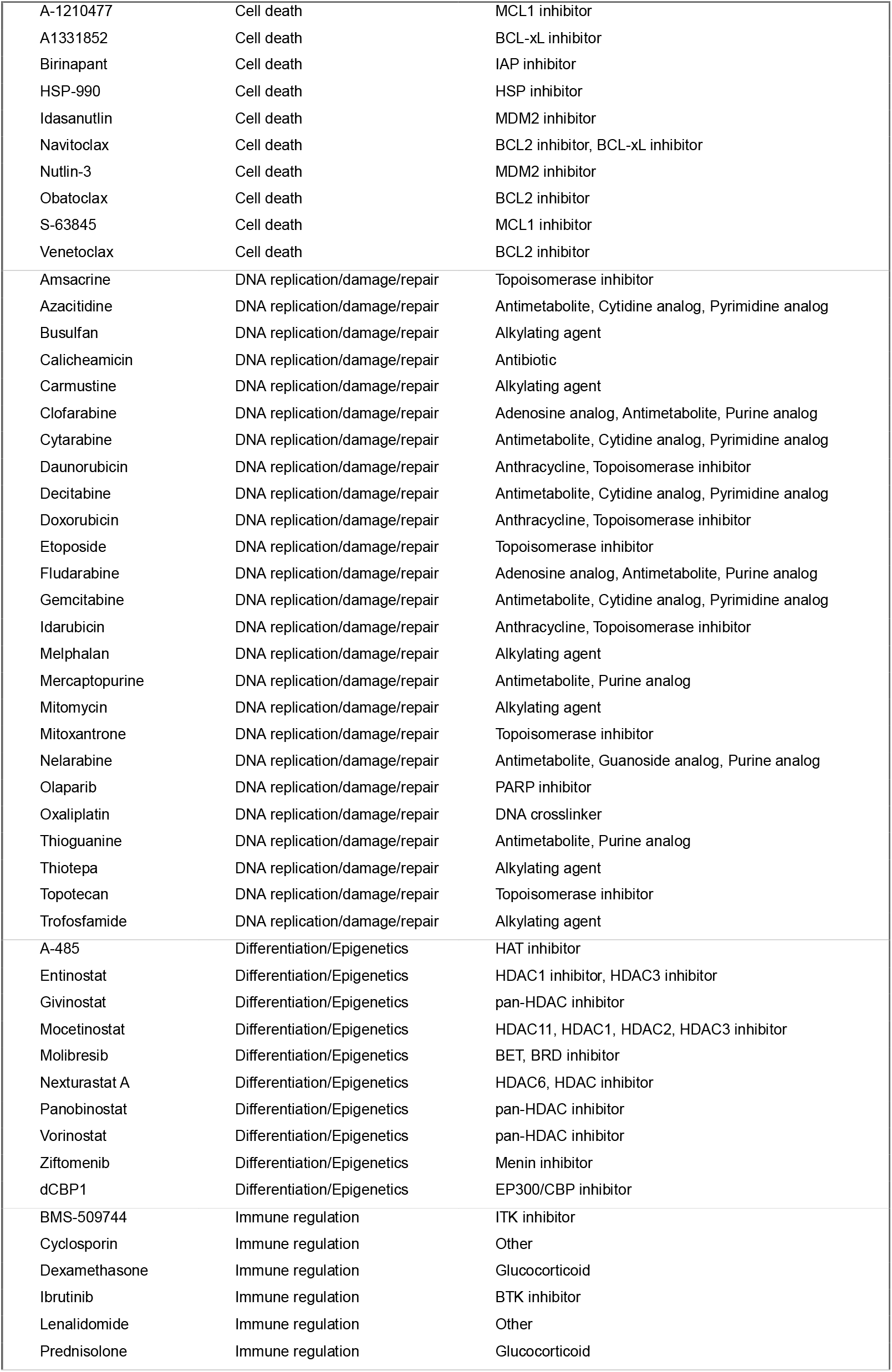
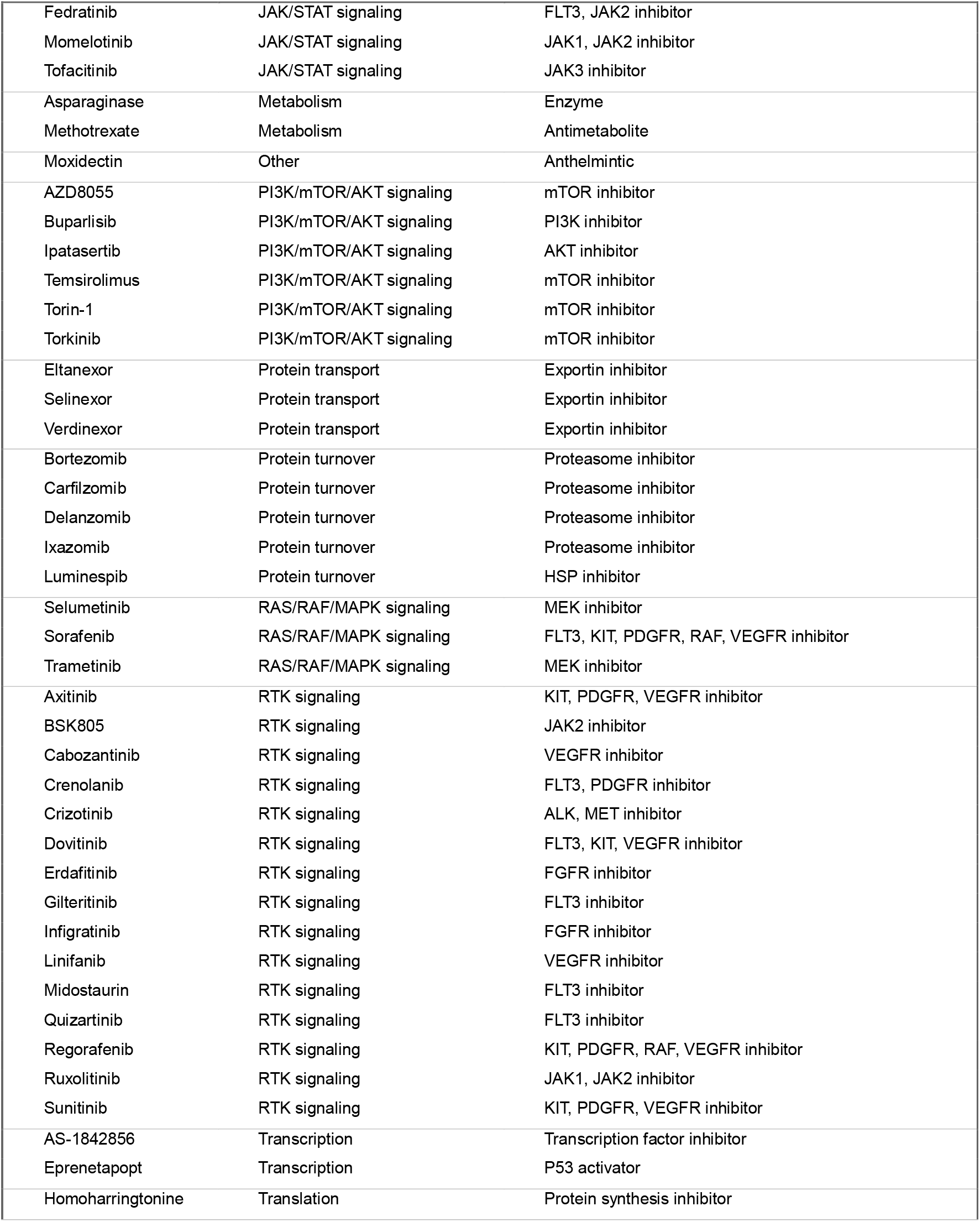
Drug library and classification.

